# Somatic mutations reveal hypermutable brains and are associated with neuropsychiatric disorders

**DOI:** 10.1101/2022.02.09.22269114

**Authors:** Taejeong Bae, Liana Fasching, Yifan Wang, Joo Heon Shin, Milovan Suvakov, Yeongjun Jang, Scott Norton, Caroline Dias, Jessica Mariani, Alexandre Jourdon, Feinan Wu, Arijit Panda, Rosalinda C. Roberts, Anita Huttner, Joel E. Kleinman, Thomas M. Hyde, Richard E. Straub, Christopher A. Walsh, Brain Somatic Mosaicism Network, Alexander Urban, James F. Leckman, Daniel R. Weinberger, Flora M. Vaccarino, Alexej Abyzov

## Abstract

Somatic mutations have causative roles in many diseases and contribute to neuropsychiatric disorders. Here, we analyzed 131 human brains (44 neurotypical, 19 with Tourette syndrome, 9 with schizophrenia, and 59 with autism) for somatic single nucleotide and structural mutations after whole genome sequencing to a depth of over 200X. Typically, brains had 20 to 60 detectable single nucleotide mutations that likely arose in early development; however, about 5% of brains harbored hundreds and up to 2000 somatic mutations. Hypermutability was associated with age and putative damaging mutations in genes previously implicated in cancer and likely reflects *in vivo* clonal expansions. Somatic duplications of likely early developmental origin were present in 1 out of 20 normal and diseased brains, reflecting background mutagenesis. Brains of individuals with autism were enriched in somatic deletions and in point mutations that create putative transcription factor binding motifs in enhancers that are active in the developing brain. The most affected motifs corresponded to MEIS transcription factors, suggesting a potential link between their involvement in gene regulation and autism.

## Introduction

Somatic mutations naturally occur in proliferative and post-mitotic cells throughout human development and during aging, starting from the first cleavage of the zygote (*1–4*). An open question is: how frequent are somatic mutations in the population, and are they a contributing factor to the etiology of neuropsychiatric disorders (*5*)? Mutations can be analyzed in single cells or in bulk tissues. In the latter case, the discovered mutations are present in a larger fraction of cells (with frequency typically above 1%), often arise during early development, and because of their higher frequency may be enriched for variants exerting a stronger phenotypic effect. Considering this premise, we investigated somatic mosaicism in normal brains (n = 44) and in the brains affected by 3 neuropsychiatric diseases: Tourette Syndrome (TS, n = 19); schizophrenia (SCZ, n = 9); and autism spectrum disorder (ASD, n = 59).

## Results

### Mutation discovery and counts across cohorts

For each frozen post-mortem brain, 1 to 2 regions (cortex, striatum, or hippocampus) were extracted (∼1 cm^3^) in one of three institutions: Yale University, the Lieber Institute for Brain Development, and Harvard University (the raw data was reported previously (*6*)). DNA extracted from bulk samples, from FACS-sorted neuronal and glial cell fractions, or from both was analyzed by whole genome sequencing (WGS) on an Illumina platform at various depths, resulting in a combined coverage above 200X per brain for 92% of the samples, and as high as 620X (**Table S1**). The data were uniformly processed – reads were aligned and somatic point mutations were called in all samples using a previously verified workflow (*7*). Somatic mutations were distinguished from inherited variations based on their frequency and based on no overlap with the catalogues of known germline variations in the human population.

In brains with bulk samples sequenced to at least 200X coverage, we discovered 20 to 60 somatic single nucleotide mutations per brain with typical allele frequencies that ranged from 1% to 10% across all cohorts (**Figs. 1** & **S1B; Table S2**). Additional mutations (tier2 mutations) were discovered when considering higher sequencing coverage including data for cell fractions (see **Methods**). Unexpectedly, in every cohort there was at least one brain that had a large burden of at least 100 somatic mutations, which we term hypermutable brain (**Fig. 1A**). The mutation calls in those brains had the same substitution spectrum as for other brains (**Fig. 1D** & **S1E**). As an additional control, we performed read-backed phasing of calls to personal DNA haplotypes in all brains. Due to the relatively short DNA fragments (∼450 bps), only ∼20% of calls were close to a nearby heterozygous SNP that could be used for phasing. The fractions of phasable and unambiguously phased calls in the hypermutable brains was the same as in other brains (**Fig. 1B**). Thus, we concluded that calls in hypermutable brains represent *bona fide* somatic mutations.

**Figure 1.**
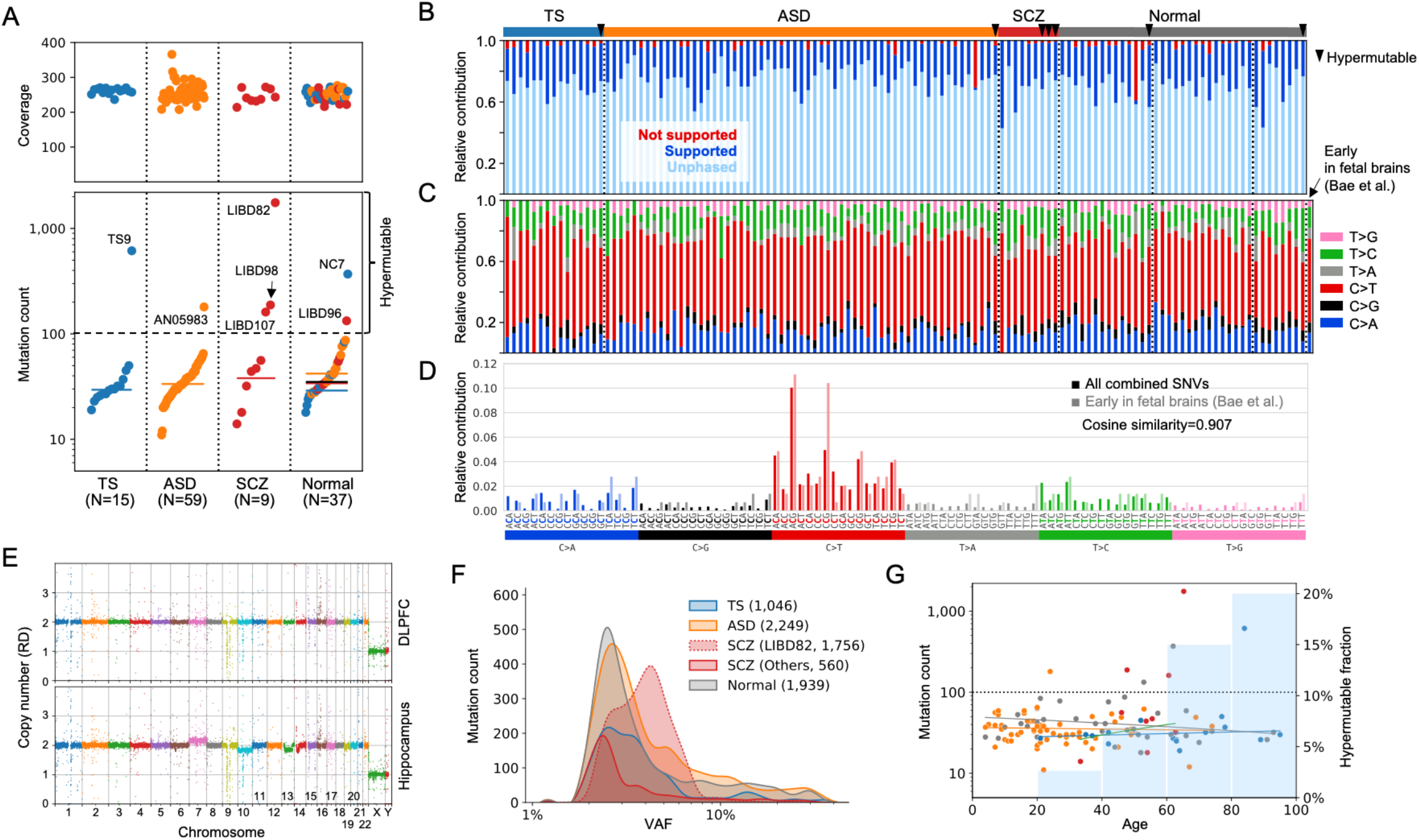
Mutations discovered across cohorts of brains. **A)** Summary of coverage and detected somatic point mutations across cohorts using bulk brain samples. Mutation burden across cohorts was comparable when excluding hypermutable brains with more than 100 of mutations (named in the plots). **B)** Fraction of supported, not supported, and unambiguously phased mutations per sample is comparable across samples (hypermutable brains are indicated by triangles). **C)** The substitution spectra of detected mutations across all brains are comparable. **D)** The combined mutation spectrum is dominated by C>T transitions in CpG motifs and matches the spectrum of developmental mutations (**Fig. S1**). **E)** Brain LIBD82 had apparent aneuploidies in the hippocampus, which may be a signature of glioma or glioblastoma, and was excluded from the downstream analyses. **F)** Distributions of mutation counts and allele frequencies across cohorts. Numbers in parenthesis list the mutation count. Brain LIBD82 is an exception and its mutations had higher frequencies than in other brains. **G)** When excluding hypermutable samples, the mutation burden does not correlate with age. However, there are more hypermutable brains (those are above the dotted line) in older brains (i.e., over 40 years old; blue bars in the background).

When we excluded the hypermutable brains, there were no significant differences in somatic mutation burden between diseased and normal brains, the allele frequency of the somatic mutations, or the somatic mutation spectrum (**Figs. 1C,D**). The combined mutation spectrum was dominated by C>T transitions in CpG motifs and matched the spectrum of mutations arising during development (**Figs. 1C,D** and **S1E**), which are thought to represent background mutagenesis in proliferative cells (*1*). Brains in each cohort had different age distributions, but the mutation burden (after excluding hypermutable samples) did not correlate with age (**Fig. S1F**), which again is consistent with somatic mutations being of early developmental origin. However, there were more hypermutable brains among older brains (over 40 years old) – about 11% – as compared to younger brains (less than 40 years old) – less than 2% (p-value = 0.04 by Fisher’s exact test) (**Fig. 1G**).

The schizophrenia cohort had a marginally higher fraction of hypermutable brains as compared to normal brains (p-value = 0.04 by Fisher’s exact test) or all other brains (p-value = 0.01). The marginal significance remained (p-value = 0.04) when we only considered brains over 40 years old. One of the hypermutable schizophrenia brains with the highest mutation burden, LIBD82, had genomic aneuploidies in hippocampus, including a duplication of chromosome 7 and a deletion of chromosome 10 (**Fig. 1E**), which was previously reported to be a signature of glioblastoma (*8*), consistent with the idea that driver mutations often precede cancer diagnosis by many years, if not decades (*9*). However, clinical characterization of the brain did not provide any evidence of the disease. Thus, hypermutability in this brain may reflect incipient glioblastoma. We excluded this brain from subsequent analyses. With that brain excluded, the enrichment of the schizophrenia cohort for hypermutable brains did not reach statistical significance.

### Origin of hypermutability

Hypermutable brains were found in patients and controls and in every cohort of samples generated by each lab. Thus, it is thus unlikely that the large mutation load in these brains is associated with disease status or arises from cohort- or lab-specific conditions used for brain storage, handling, or sample processing. Thus, we hypothesize that the hypermutability arises from individual-specific biological causes. One possibility is that cells of such brains are intrinsically hypermutable. However, the frequency spectrum of mutations in hypermutable as compared to non-hypermutable brains was depleted for higher frequency mutations (>6% VAF), strongly overrepresented for intermediate frequencies (3%-6%), and almost the same for low frequency (<3% VAF) mutations (**Figs. S1B-D**). These data suggest that intrinsic hypermutability, if it exists, is not innate (that is, present in all cell divisions throughout development), but occurred sometime during the lifetime of that individual.

Out of 6 hypermutable brains, two, TS9 and NC7, had the same known missense mutation (chr1:115,258,747 C>T) in the *NRAS* oncogene. This mutation has been previously identified in multiple cancers and is deemed to be pathogenic in the COSMIC database (ID: COSV54736383). Furthermore, 3 brains (NC7, TS9, and AN05983) carried 11 mutations predicted to have a damaging effect in genes previously implicated in cancers (**Table 1**), such as *MTOR, TET2, DNMT3A*, and *IDH2*. Some of the mutations were also annotated in the COSMIC database as being pathogenic, and one more (besides the mutation in NRAS) pathogenic mutation in 3’-UTR of *HNRNPA2B1* was recurrent. Hypermutable brains were overall enriched for damaging mutations in genes implicated in cancers (*11, 12*) as compared to non-hypermutable brains (p-value = 2.4×10^−3^ by Fischer’s exact test; **Fig. S2A**).

**Table 1.** List of detected somatic mutations that putatively affect genes previously implicated in cancers and genes previously associated with neuropsychiatric diseases. Some mutations are also annotated as pathogenetic in the COSMIC database (*10*). A known mutation in *NRAS* oncogene is present in two brains. Additionally, a 3’-UTR mutation (chr7:26,231,874 A>T) in the cancer implicated gene *HNRNPA2B1* was present in two brains and was included in the table for that reason. *tier2 mutation (see **Methods**); Validation: **A – by amplicon, R – previously reported and validated in Rodin et al. (*6*), S – by single cells; Gene annotation: ***B – cancer implicated genes in Bailey et al. (*11*), C – cancer gene census (*12*), ASD – SFARI genes, SCZ – compiled from previous SCZ studies (*13–16*).

| Mutation | Brain | Phenotype | Hypermutable | VAF | Validation** | Consequence (VEP) | Gene | Gene annotation*** | Genomic Mutation ID (COSMIC) | Prediction (COSMIC) |
| --- | --- | --- | --- | --- | --- | --- | --- | --- | --- | --- |
| **In genes implicated in cancer** |  |  |  |  |  |  |  |  |  |  |
| chr1:115,258,747 C>T | NC7 | Normal | Yes | 1.84% | A, S | Missense | *NRAS* | B, C | COSV54736383 | Pathogenic |
|  | TS9 | TS | Yes | 1.10% | A |  |  |  |  |  |
| chr2:25,466,766 C>G | NC7 | Normal | Yes | 1.91% | A, S | Splice | *DNMT3A* | B, C | - | - |
| chr3:81,695,552 G>T | TS9 | TS | Yes | 1.26% | n/a | Missense | *GBE1* | - | COSV101450116 | Pathogenic |
| chr4:106,156,747 C>T* | TS9 | TS | Yes | 1.05% | A | Stop | *TET2* | B, C | COSV54395664 | Neutral |
| chr4:106,197,392 G>T |  |  | Yes | 1.75% | A | Stop |  |  | - | - |
| chr4:183,267,873 C>T | TS9 | TS | Yes | 1.27% | n/a | Missense | *TENM3* | - | COSV69301741 | Pathogenic |
| chr12:48,107,157 C>G | TS9 | TS | Yes | 1.16% | n/a | Missense | *ENDOU* | - | COSV57465821 | Pathogenic |
| chr15:90,631,934 C>T* | NC7 | Normal | Yes | 3.55% | S | Missense | *IDH2* | B, C | COSV57468751 | Pathogenic |
| chrX:129,168,460 C>T | NC7 | Normal | Yes | 2.99% | S | Stop | *BCORL1* | C | COSV54390206 | Neutral |
| chr1:11,300,357 T>C | AN05983 | ASD | Yes | 2.63% | A | Splice | *MTOR* | B, C | - | - |
| chr7:26,231,874 A>T | LIBD96 | Normal | Yes | 4.52% | n/a | 3' UTR | *HNRNPA2B1* | C | COSV61161436 | Pathogenic |
|  | LIBD99 | Normal | No | 10.28% | n/a |  |  |  |  |  |
| chr10:32,326,503 A>T* | LIBD99 | Normal | No | 2.62% | n/a | Missense | *KIF5B* | C | - | - |
| chr6:168,315,889 C>T* | TS1 | TS | No | 0.95% | n/a | Stop | *MLLT4* | C | COSV60045383 | Pathogenic |
| chr4:1,977,081 G>A | UMB797 | ASD | No | 2.78% | n/a | Missense | *WHSC1* | B, C | COSV56389769 | Pathogenic |
| chr7:148,495,674 A>G | UMB5297 | ASD | No | 2.99% | n/a | Missense | *CUL1* | B | - | - |
| **In disease-implicated genes in brains with diseases** |  |  |  |  |  |  |  |  |  |  |
| chr1:11,300,357 T>C | AN05983 | ASD | Yes | 2.63% | A | Splice | *MTOR* | ASD | - | - |
| chr2:179,612,387 T>A | UMB1638 | ASD | No | 2.09% | n/a | Stop | *TTN* | ASD | - | - |
| chr8:146,033,199 C>T | AN13654 | ASD | No | 4.15% | n/a | Missense | *ZNF517* | ASD | - | - |
| chr10:55,600,099 C>T | UMB4231 | ASD | No | 4.41% | A | Missense | *PCDH15* | ASD | COSV57387920 | Pathogenic |
| chr16:20,976,490 C>T | AN09412 | ASD | No | 12.09% | R | Missense | *DNAH3* | ASD | - | - |
| chr2:73,800,041 T>G* | LIBD100 | SCZ | No | 4.46% | n/a | Missense | *ALMS1* | SCZ | - | - |

Given the above data, we hypothesized that the cell lineage carrying one or more of these mutations underwent a clonal expansion in the brain, which allowed other preceding lineage-specific mutations to arise to a detectable frequency. In both TS9 and NC7 brains, virtually all of the mutations were present in both bulk cortex and striatum at a frequency of about 2%, as well as in multiple other cell fractions at various frequencies (**Figs. 2A,B** and **S2B,C**). Furthermore, in brain NC7, all the mutations were present in sorted interneurons from the striatum (STR-INT; see **Methods**) at a much higher frequency (15-25% VAF), suggesting that in this brain, the expanded lineage originated in the embryonic basal ganglia, where interneurons are generated, and then populated cortex and striatum by cell migration.

**Figure 2.**
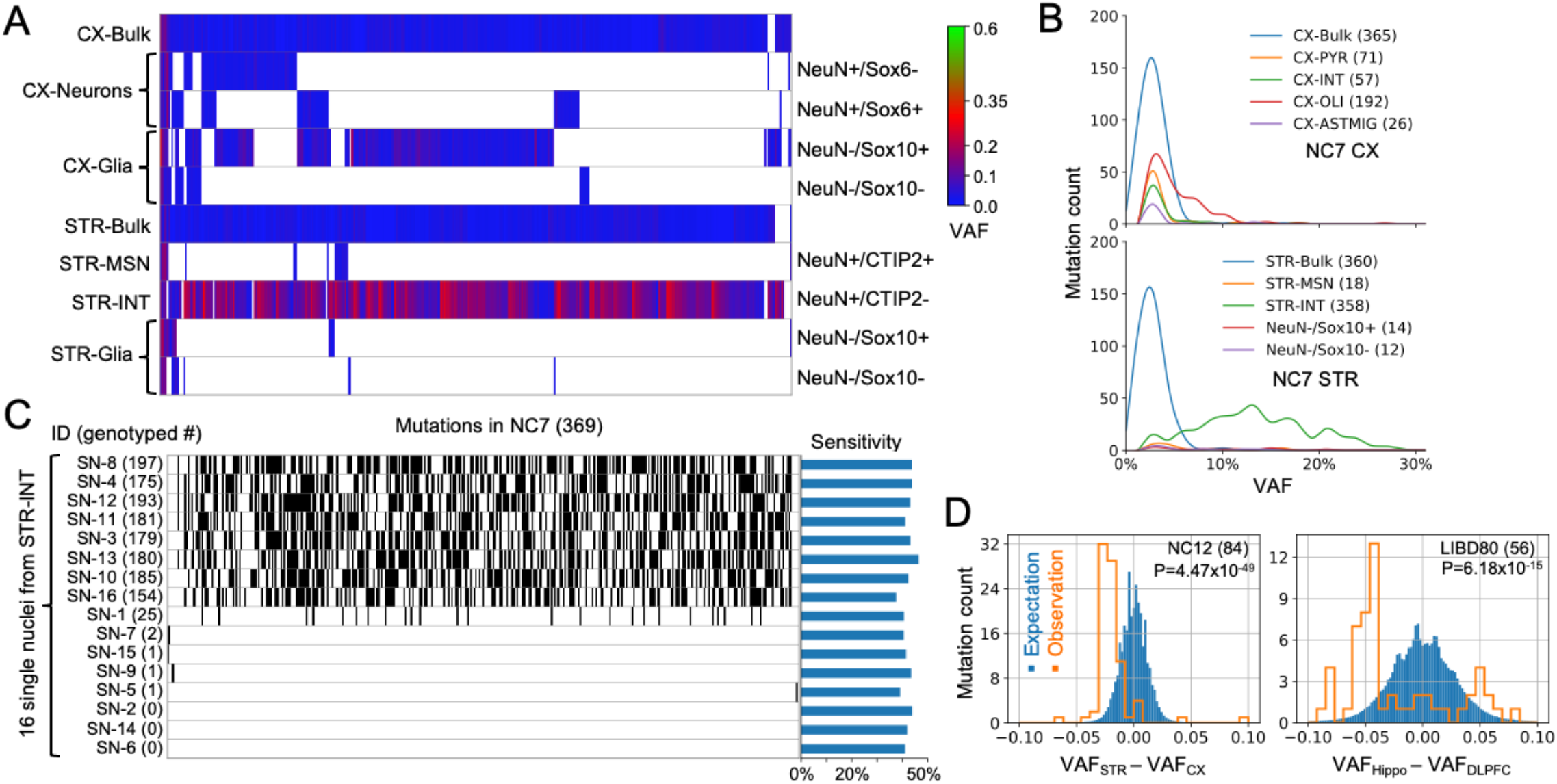
Uneven cell lineage distribution of somatic mutations in brain. **A)** Mutation allele frequencies in samples from brain NC7. Almost all mutations discovered in NC7 are present in the striatal interneuron fraction (STR-INT) at high VAFs. **B)** Distribution of mutation allele frequencies across samples in NC7. **C)** Genotyping of mutations in 16 single nuclei originated from the NC7 STR-INT fraction. Black bars represent genotyped mutations. **D)** The frequencies of mutations are significantly biased toward cortex as compared to striatum (NC12) or as compared to hippocampus (LIBD80).

We tested the above inference by sequencing (at 5X coverage) 16 single nuclei isolated from a different sample of the STR-INT fraction from NC7. Of all discovered somatic mutations in bulk NC7 brain, 346 (94%) were genotyped and validated in eight out of 16 nuclei, with 154 to 197 mutations genotyped in each nucleus (**Fig. 2C**). The genotyping rate, 42% to 53% across nuclei, was comparable to the genotyping sensitivity (34% to 46%, see **Methods**) allowing us to extrapolate that each of the eight nuclei contained almost all the mutations discovered in the bulk cortex and striatum, and proving that the mutations belonged to the same cell lineage. Differently than NC7 and TS9, mutations in the three other hypermutable brains with data available for multiple regions (LIBD96, LIBD98, and LIBD107) were generally restricted to one brain region (**Fig. S2D**); thus, hypermutability in the latter brain samples was more consistent with a local lineage expansion, perhaps later in life.

### Somatic Mutations revealed uneven cell lineage distribution in brain

For each brain processed at Yale University (19 TS and 21 neurotypical brains), we obtained sequencing data for bulk cortex, striatum, and for up to 8 cell fractions per brain region (see **Methods**). In both the cortex and striatum, we successfully fractionated neurons and non-neuronal cells (NeuN+/NeuN-); in the striatum, we also fractionated medium spiny neurons (STR-MSN) and interneurons (STR-INT) (**Fig. S22** and **Methods**). The frequency of mosaic mutations in multiple regions and fractions allowed us to explore the cell lineage distribution in the brain. For example, as described above, almost all the somatic mutations discovered in the hypermutable brain NC7 were found in the STR-INT fraction at higher VAF than in the bulk tissues, suggesting interneuron lineage expansion in the striatum (**Figs. 2A,B**).

To investigate whether the above result could reflect a more general phenomenon, we tested for the difference in mutation VAFs between the cortex and striatum. If the early lineage distribution is the same across these two regions, there should be, on average, no difference in VAFs between them (see **Methods**). Out of 22 brains of the Yale cohort for which we had a complete dataset for the bulks and fractions, eight (six non-hypermutable) showed a statistically significant difference between the cortex and striatum (**Figs. 2D** and **S3A,B**). Except for the NC7 hypermutable brain, a higher VAF of somatic mutations was always observed in the cortex. Even in NC7, the VAFs were higher in cortex when excluding neuronal fractions (**Fig. 2A, S3C**). Similarly, in 5 out of 13 non-hypermutable brains processed by the Lieber Institute (6 SCZ and 7 neurotypical brains), we also observed a higher VAF in cortex as compared to the hippocampus (**Figs. 2D** and **S3D,E**). Combining the brains with biased VAFs from the two datasets, we observed a significant overrepresentation of brains with higher VAF values in the cortex (p-value = 5×10^−4^ by binomial test). A possible explanation of this observation is that the founder population of the cortex is allocated with fewer earlier lineages, so that the frequency of each lineage is, on average, higher. Alternatively, the cortex may have a higher propensity for expansion in some lineages, when compared to the striatum and hippocampus.

### Impact of somatic mutations on genome function

Given the mostly random distribution of somatic mutations in the human genome, the majority (∼60%) of the mutations were outside genes with roughly the same distribution across the genome in each cohort (**Figs. 3A** and **S4A**). Approximately 2% to 3% of mutations were in the coding part of the genome and another 6% had the potential to affect regulatory regions. When considering the mutations affecting gene functions, 6 putative deleterious mutations in genes previously associated with neuropsychiatric diseases were found in ASD and SCZ brains (**Table 1**). For example, in the UMB4231 ASD brain, we detected a missense mutation in *PCDH15* – a gene encoding a protein mediating calcium-dependent cell-cell adhesion, which was previously associated with ASD (*17, 18*). Furthermore, in the AN05983 ASD brain, we detected and validated a splice mutation in *MTOR*, a gene which is part of the IGF1/PI3K signaling pathway that previously was implicated in ASD (*19–21*).

**Figure 3.**
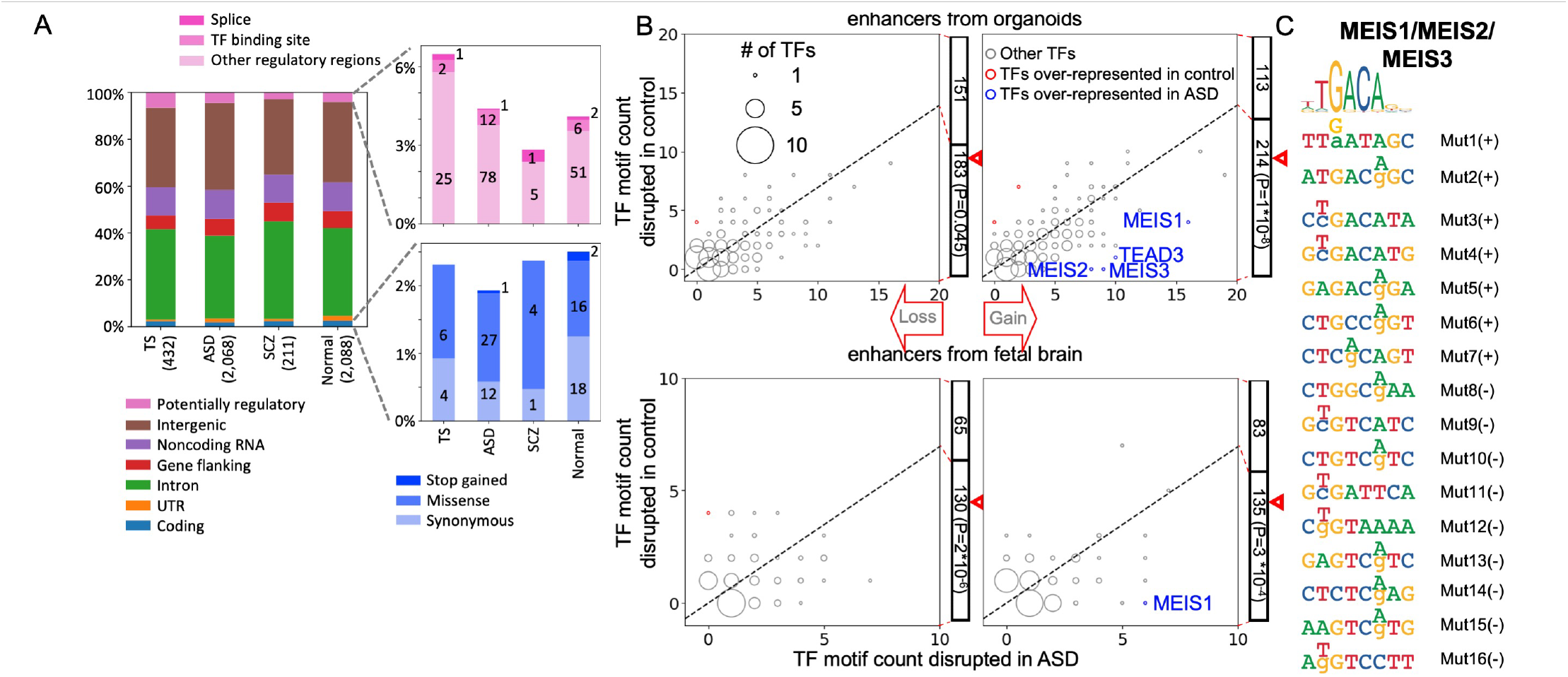
The impact of somatic point mutations on genome function. **A)** In each cohort mutations from non-hypermutable brains have roughly the same distribution across genomes and similar predicted functional impacts by VEP. **B)** Counts of putative TF motifs disrupted by somatic mutations within enhancers regions from organoids (upper plots) or fetal brain (lower plots), showing that mutations significantly disrupt putative TF binding sites in ASD vs controls. The size of the dots represents the number of TFs with the same ASD and control counts. **C)** Somatic mutations within putative enhancer regions resulting in gain of binding sites for MEIS1, MEIS2, and MEIS3. The consensus motif is on top. Mutations on positive (+) or negative (-) strand are enumerated on the right with coordinates listed in **Table S3**. For each mutation, a reference base is show in small letter and the mutation base is noted by a capital letter above.

While we did not detect deleterious somatic mutations in TS associated genes, a missense mutation in the *ARHGEF6* gene (chrX:135,761,734 G>T) was found in brain TS9 at a relatively high VAF in both the cortex (4.2% VAF) and striatum (1.6% VAF). The *ARHGEF6* gene has been associated with X-linked intellectual disability (*22, 23*) and dendrite orientation/cell polarity (*24*). Indeed, knockout of *ARHGEF6* in mouse models results in hippocampal dendrite/synaptic abnormalities and deficits in learning (*25*). Intriguingly, deficits in cell polarity have been implicated in Tourette syndrome from analyses of rare *de novo* mutations (*26*).

We next investigated the functional impact of the discovered mutations on enhancers active in the fetal human brain and in an organoid model of developing human brain (see **Methods**). For each set of enhancers, analysis of the ASD cohort revealed an excess, as compared to controls, of somatic mutations creating putative transcription factor binding motifs (p-value < 10^−4^ by binomial test) (**Fig. 3B** and **S4C**). Most prominently, 15 putative binding motifs for the MEIS1, MEIS2 and MEIS3 transcription factors were created by mutations in ASD brains, whereas only 3 of such mutations were detected in normal brains (**Fig. 3C**).

### Somatic structural mutations

To call structural mutations, we used an approach implemented in CNVpytor (**Fig. 4**). The tool segments the genome, considering both depth of coverage and split in allele frequencies for single nucleotide polymorphisms (SNPs), allowing us to call deletions, duplications, and copy number neutral losses of heterozygosity (CNN-LOH). The called mutations can be either inherited or somatic and we distinguished between these two types by their cell frequency (**Methods**). Almost half of the structural mutations were validated by mapping their exact breakpoints. For the remaining mutations, we imputed haplotypes and confirmed them by calculating the imbalanced frequency of the haplotypes (**Table S4**; **Figs. S5-S18**). For two mutations, we attempted validation in brain tissue by PCR followed by amplicon-seq across the breakpoints, which confirmed them both (**Figs. 4** and **S5**).

**Figure 4.**
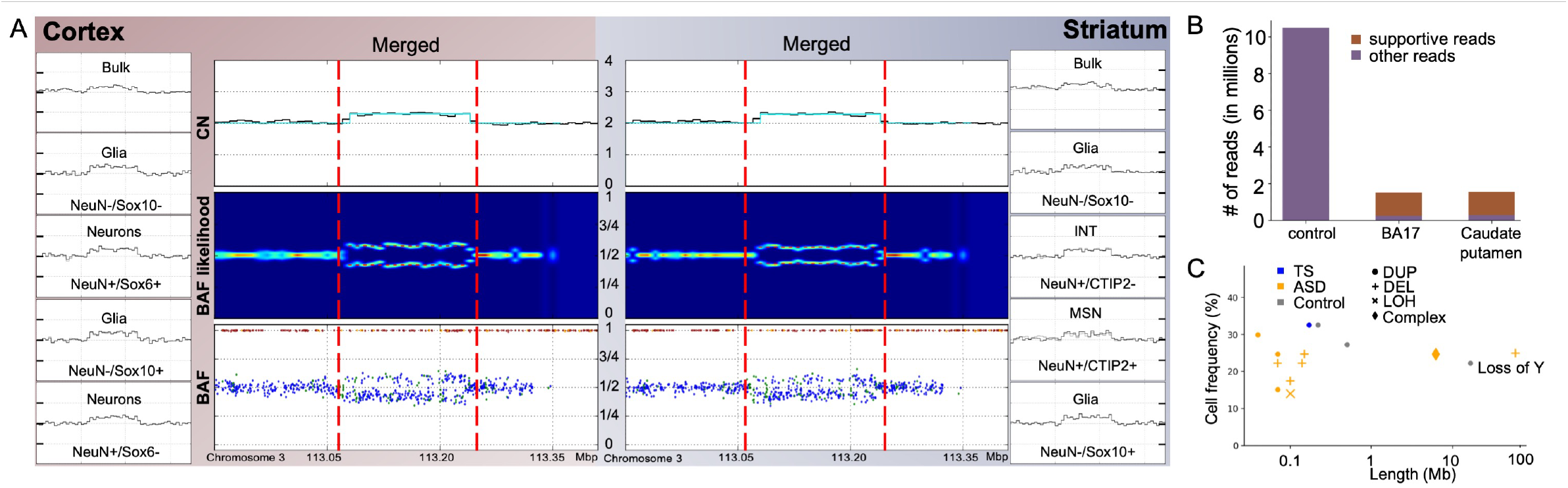
Detection and validation of somatic large structural mutation in brain TS1. **A)** Example of a duplication on chromosome 3 (chr3:113,067,261-113,233,476) identified in brain TS1 in eight cell fractions and two bulks: cortex and striatum. Red vertical lines outline the region of the duplication event. The two combined plots for cortex (left) and striatum (right) present the read depth (top), VAF likelihood function (middle) and BAF for individual SNPs (bottom) calculated from combined fractions and bulk data. SNPs in the accessible genome (P-bases) are in blue, other SNPs are in green. **B)** Amplicon-seq validation of the duplication in A). Bar plot show the number of reads mapping to the junction of the duplication in the cortical region BA17 and caudate putamen of TS1 and in control sample. **C)** Summary of size and cell frequency of all structural mutations detected from all analyzed individuals.

In total, we detected 14 rearrangements in 13 (10%) brains (all non-hypermutable) ranging from 40 kbp to 16.5 mbp in length, with the largest mutation representing the loss of chrY. Most somatic structural mutations were duplications, which were found in about 5% normal and diseased brain. The two duplications discovered in the Tourette cohort (brain TS1 and NC6) were present in all bulk samples and fractions analyzed (**Fig. 4** and **S5**). This finding is consistent with the idea that these duplications occurred during development, which is also supported by their frequency in at least 10% of cells. Consistent with a developmental origin, sequence microhomologies (1-4 bps) were found in all but one of the resolved breakpoints (**Fig. S20**), suggesting that the corresponding mutations were generated by replicative mechanisms, as previously observed for mosaic CNA in the fetal human brain (*27*). In many aspects, duplications resemble *de novo* CNVs in the CNV Mutator Phenotype (*28*) – in that they are mostly tandem duplications of early developmental origin and have sequence microhomologies at breakpoints – although the mutations detected here were smaller than reported in previous studies. Thus, we hypothesize that somatic duplications represent background mutations that arise in early development, with the CNV mutator phenotype representing the extreme manifestation of this phenomenon leading to a disease phenotype. Mutations where breakpoints could not be resolved may have arisen from DNA repair mechanisms relying on long sequence homologies and repeats around breakpoints, which make the breakpoints inaccessible to short read sequencing.

Remarkably, the ASD cohort was the only one where deletions and CNN-LOH mutations were discovered and significantly overrepresented when compared to all other brains (p-value = 0.008 by Fischer’s exact test). Additionally, one deletion and one complex rearrangement (but mostly containing duplicated sequence) occurred in regions where previous studies associated germline CNVs with an ASD phenotype (*29–32*) (**Table S5**). For example, in the AN14067 brain, the deletion occurred in the ASD associated 16p13.3 region and deleted the *SOX8* and *LMF1* genes (*29, 30*). Overall, these findings point to the relevance of somatic structural mutations to the ASD phenotype.

## Discussion

Our study revealed a complex etiology of somatic mutations in brain and their possible relationship with phenotype. While most of the brains had just a few dozen mutations of likely developmental origin in proliferating cells, a significant fraction (∼6%) of brains were classified as hypermutable. Hypermutability was not related to disease, but increased with age, reaching at least a 3% population frequency (95% confidence interval) for brains over 40 years old. In some brains hypermutability could be related to clonal gliogenesis in adult brains, consistent with previous reports of increased glial fraction with age (*33, 34*). Half of the hypermutable brains carried mutations in genes that have been involved in clonal hematopoiesis in aging adults (*35–37*), consistent with previous observations from the analysis of a panel of genes (*38*). Thus, it is tempting to speculate that increases in clonal hematopoiesis with age (*37*), coupled with a leakier blood-brain barrier with age and extravasation of blood in brain tissue (*39*), could explain the higher mutation burden in those brains. However, in one of such brains, NC7, all mutations were present in two brain regions and were enriched in post-mitotic neuronal cell fractions, giving support to yet another possibility, that of a neural lineage clonal expansion during development. In support of this scenario, a spectrum of *de novo* variants in *DNMT3A*, which overlaps with those found in hematologic malignancies, causes Tatton-Brown-Rahman syndrome, a developmental disorder with macrocephaly (*40*). Neural lineage clonal expansion during development is potentially a new and important factor that could influence human phenotypes. Nonetheless, firmly establishing the cause(s) and extent of the phenomenon of hypermutability requires analysis of larger sets of brains, and additional analyses at the single cell level.

We detected associations between somatic mutations and phenotype only in the ASD cohort. This could reflect the fact that the ASD cohort was the largest, and we suggest that analyses of larger cohorts of brains with TS and SCZ phenotypes is necessary to exclude a possible association of somatic mutations with those phenotypes. The association of large and *de novo* germline CNVs (mostly deletions) with ASD is known (*41*) as the most frequent genomic factor contributing to the disorder (*42*). Furthermore, developmental disorders and ASD were associated with somatic CNVs in the blood (*43, 44*). Our study reveals that a similar association exists for somatic deletions and CNN-LOH in the brain, suggesting the importance of assessing somatic structural mutations for understanding the pathophysiology of ASD.

In ASD brains, somatic point mutations within noncoding regions created putative TFs binding motifs within enhancers that are active in the developing human brain. We hypothesize that some of the newly created motifs may bind their corresponding TFs, thereby affecting TF protein dosage and dysregulating gene regulatory networks. The top affected motifs are putative binding sites for MEIS genes, which are homeodomain-containing transcriptional activators that promote chromatin decompaction (*45*). MEIS TFs have been implicated in intellectual disabilities, act as cofactors of HOX genes, and are important regulators of proliferation, growth, neurogenesis and patterning during development (*45, 46*). Interestingly, MEIS2 expression identifies a distinct subpopulation of cortical interneurons that populates the white matter (*47*), highlighting its potential importance in ASD. Using a brain organoid model, we previously proposed that ASD deleterious *de novo* variants are significantly enriched in homeodomain TF binding motifs within developmental enhancers (*48*). Similarly, another study of brain development using organoids found that enhancers associated with ASD genes from the SFARI collection are enriched for homeobox binding motifs (*49*). Therefore, we suggest that functional studies of mutations within homeodomain binding motifs during human development represent an important direction of future experimental efforts in understanding the etiology of ASD.

## Data Availability

All data produced in the present study are available in NIMH Data Archive.

https://nda.nih.gov/study.html?id=814

## Availability of data

Data and call sets have been deposited in the NIMH Data Archive (NDA Study ID 814) and can be accessed as part of the NIMH Data Archive permission groups: https://nda.nih.gov/user/dashboard/data_permissions.html.

## Acknowledgments

The authors thank the Tourette’s Association of America for collecting and providing brain tissue. We also thank the donors and their families. We acknowledge the NIH Neurobiobank and its Director, Dr. Sabina Berretta, for providing postmortem tissue from TS and NC individuals. We are grateful to Geoff Lyon of the Yale Flow Cytometry Facility for help with FACS sorting of brain postmortem nuclei. We thank member of BSMN consortium for useful discussions and suggestions during the study and preparation of the manuscript. Members of the BSMN consortiums are listed in the supplementary file. We express our gratitude to the NDA and Synapse team for providing collaborative data storage space and support for data transfer needed for the successful completion of this study. This study was supported by NIH grants U24CA220242, U01MH106892, U01MH106876, U01MH106883, U01MH106874, U01MH106893, U01MH106882, U01MH108898, U01MH106891, U01MH106884, and R01MH109648; and by SFARI grant 399558.

## Ethics Statement

The analyzed samples and generated data were de-identified and derived from an individual postmortem.

Brains with Tourette Syndrome sequenced by Yale University were obtained from the Harvard’s Brain and Tissue Resource Center (HBTRC). Normal brains sequenced by Yale University were obtained from NIH NeuroBioBank. The work with those brains was handled in multiple institutes under multiple IRBs:

### University of Miami Brain Endowment Bank

IRB Protocol Number: 19920358 (CR0001775)

- IRB: University of Miami Institutional Review Board
- Title: Brain Endowment Bank

### University of Maryland Brain and Tissue Bank

IRB Protocol Number: HM-HP-00042077

- IRB: The University of Maryland Institutional Review Board
- Title: University of Maryland Brain and Tissue Bank

IRB Protocol Number: 5-58

- IRB: The Maryland Department of Health and Mental Hygiene IRB
- Title: University of Maryland Brain and Tissue Bank

### Harvard Brain Tissue Resource Center

IRB Protocol Number: 2015P002028

- IRB: Partners Human Research Committee
- Title: Brain Tissue Repository for Research on Neurological and Psychiatric Disorders

### The Human Brain and Spinal Fluid Resource Center (managed by Sepulveda Research Corporation)

IRB Protocol Number: PCC#: 2015-060672, VA Project #: 0002

- IRB: Department of Veterans Affairs - Los Angeles
- Title: NIH Brain and Tissue Repository - California

### Mt. Sinai Brain Bank

IRB Protocol Number: HAR-13-059

- IRB: Bronx VA Medical Center Institutional Review Board
- Title: The NIMH, NICHD, NINDS Brain and Tissue Repository

### Brain Tissue Donation Program at the University of Pittsburgh

IRB Protocol Number: REN14120157/IRB 981146

- IRB: University of Pittsburgh Institutional Review Board
- Title: University of Pittsburgh NIMH, NICHD, NINDS Brain and Tissue Repository

For normal brains from Yale University autopsy authorization, consent to utilize tissues for research and consent to publish was provided by the patient’s next-of-kin according to Connecticut state law and approved protocols of the Yale University BioBank. The research study was approved by the Yale Alzheimer Disease Research Center (ADRC) and was reviewed and deemed exempt by the Yale University Institutional Review Board. The BioBank protocols are in accordance with the ethical standards of Yale University.

Handling of brains originating from and sequenced by the Lieber Institute was conducted under the approved WCG IRB protocol 20111080 titled “Collection of Postmortem Human Brain, Blood and Scalp Samples for Neuropsychiatric Research”.

Data for brains sequenced by Harvard University were publicly available from NIMH Data Archive.

## Supplementary Materials

### Material and Methods

#### Preparation and sequencing of samples by Yale (contributed by Liana Fasching)

Brains of Tourette’s patients were dissected and stored as per Harvard’s protocol (*50*). Dr. Roberts visited the HBTRC at Harvard and dissected approximately 1cm chunks of frozen brain tissue for this study.

#### Isolation of nuclei and fractionation of bulk tissue

Per sample, 1.5g of postmortem tissue (CX BA6 or caudate nucleus and putamen of the striatum (STR)) was dissected into 2 mm cubes. Tissue was lysed and nuclei were extracted following the protocol described by Matevossian et al. (*51*). Nuclei were resuspended in ice-cold PBS and BSA (dilution 1:100). Nuclei isolated from caudate nucleus and putamen (striatum), were incubated with the following combination of antibodies: anti-NeuN conjugated to 488 fluorophores (Millipore clone A60, MAB377X; 1:500) to sort for NeuN+ neurons versus NeuN-glia and anti-CTIP2 conjugated to 647 fluorophores (Abcam ab18465, 1:250; conjugation in house with Alexa Fluor^™^ 647 Antibody Labeling Kit, Thermo Fisher, A20186) to be able to separate medium spiny neurons (NeuN+/CTIP2+) from striatal interneurons (NeuN+/CTIP2-) (*52*). Simultaneously, these nuclei were incubated with anti-Sox10 antibody (Cell Signaling, 1:500) and secondary antibody Cy^™^3 AffiniPure Donkey Anti-Rabbit IgG (H+L) (Jackson Laboratory, 711-165-153; 1:200) to isolate Sox10+ oligodendrocytes (NeuN-/CTIP2-/SOX10+), while the triple negative fraction (NeuN-/CTIP2-/Sox10-) was likely enriched with astrocytes and microglia. For the cerebral cortex, since cortical interneurons express Sox6, we used an anti-Sox6 antibody conjugated to 647 fluorophore antibody (Abcam ab30455, 1:500; conjugation in house with Alexa Fluor^™^ 647 Antibody Labeling Kit, Thermo Fisher, A20186) together with NeuN to obtain pyramidal neuron (NeuN+/Sox6-), and interneuron fractions (NeuN+/Sox6+), while we used the same antibodies used in the striatum to isolate oligodendrocytes and astrocyte fractions (**Fig. S21**).

Hierarchical clustering of transcriptomes from extracted fractions validated the separation of neurons from glia in both cortex and striatum using NeuN (**Fig. S22**). Similarly, CTIP2 was effective in striatum for separating medium spiny neurons (STR-MSN with NeuN+/CTIP+) from interneurons (STR-INT with NeuN+/CTIP-). The SOX6 and SOX10 antibodies weren’t as effective in separating cortical interneurons from pyramidal neurons and oligodendrocytes from astrocytes, therefore the exact correspondence of cells type to the derived fraction is not known.

Nuclei were sorted as fractions into lysis buffer (DNeasy Blood and Tissue Kit (QIAGEN)) by Fluorescent Activated Nuclei Sorting (FANS) (BD FACSAria™ II). DNA was extracted using the DNeasy Blood and Tissue Kit (QIAGEN) including an RNAse A treatment step according to suppliers’ recommendations with adjustments in proportion to volume. DNA was eluted in 100 μl elution buffer and the DNA concentration was measured by Qubit (ThermoFisher Scientific).

#### Library preparation and WGS of nuclear fractions from postmortem brain tissue

Library preparation (240 fractions: TruSeq PCR-Free 450 bp, 500ng DNA input, 32 fractions: TruSeq Nano, 450bp, 200ng DNA input) and WGS was performed by NYGC and sequenced under following conditions: Illumina HiSeqX, 2×150bp reads, 30X coverage.

#### DNA isolation and WGS of bulk postmortem brain tissue

0.1g of bulk tissue was incubated at 56°C overnight in lysis buffer (Quiagen) and DNA was extracted according to suppliers’ recommendations including RNAse A treatment. DNA was eluted in 200 μl elution buffer and the DNA concentration was measured by Qubit (ThermoFisher Scientific). Library preparation: Macrogen (library: Illumina Truseq DNA PCR-free (350bp insert), DNA input: 1μg) or NYGC (library: TruSeq PCR-free, 450bp, DNA input: 800ng) and WGS was performed by either Macrogen (Illumina, 2×150bp, 100x) or NYGC (Illumina HiSeqX, 2×150bp, 100 x coverage).

#### Preparation and sequencing of samples by the Lieber Institute for Brain Development (LIBD)

(contributed by Richard Straub)

A series of matching procedures on case and control brain samples was performed to maximize the likelihood of finding genomic mosaicism associated with diagnosis. Pairs were matched on sex, ancestry, and age at death, and various confounders that compromise molecular studies of postmortem human brain controlled for. Samples that had fibroblast cell lines available were included and those with evidence of known recurrent CNVs associated with schizophrenia were excluded.

#### DNA extraction

Genomic DNA from dorsolateral prefrontal cortex (DLPFC) and hippocampus was extracted from 100mg pulverized grey matter by Qiagen AllPrep DNA/RNA/miRNA Universal Kit (Cat#80224) or QIAamp Fast DNA Tissue Kit (Cat#51404) according to manufacturer’s instructions. DNA concentration was measured by Qubit (ThermoFisher Scientific).

#### WGS library preparation and sequencing

Illumina TruSeq Nano DNA kit based on multiple QC metrics. Each library was sequenced at 90x coverage at Psomagen (Rockville, MD) using a NovaSeq 6000 S4 flowcell with 150bp paired-end reads. DNA input for library preparation was 300ng for bulk WGS and 30ng DNA for Prime Flow RNA Assay WGS.

#### Alignment of sequencing data (contributed by Yeongjun Jang and Taejeong Bae)

We used the genome mapping command in the BSMN pipeline (https://github.com/bsmn/bsmn-pipeline) for the uniform processing of sequencing data. It implements the best practice protocol suggested by GATK described as follows. First, we split the original FASTQ files into separate files per flow cell lane. If original FASTQ files were not available, we converted original BAM files to FASTQ format. Then, we separately aligned them per lane to the human reference genome GRCh37d5 (ftp://ftp-trace.ncbi.nih.gov/1000genomes/ftp/technical/reference/phase2_reference_assembly_sequence/hs37d5.fa.gz) using bwa (version 0.7.17) so that reads from the same flow cell lane became the same read group. Next, aligned BAM files were sorted per read group and merged into a single BAM file with sambamba (version 0.6.7). Then, we removed duplicate reads using the mark duplicate command of PICARD (version 2.12.4). Finally, we performed indel realignment and base quality recalibration using GATK3 (version 3.7), resulting in the final uniform-processed BAM files.

#### Calling of somatic point mutations (contributed by Taejeong Bae)

Calling of somatic point mutations was performed following a single sample approach described in the BSMN reference brain study (*7*). Briefly, we ran GATK4 (version 4.1.2) HaplotypeCaller with the ploidy value of 50 to get raw call sets. Then, we applied the first seven filtering steps described in the above study to obtain high-quality candidates. We additionally filtered out the sites within the region of more than six short homo-, di-, and tri-polymeric tandem repeats. Finally, we applied MosaicForecast to select final call sets. We separated the calls into high confidence calls (mosaic_P by MosaicForecast > 0.6) and extended calls (remaining).

We designed different combined calling strategy per data as each data have different sample composition and sequencing coverage (**Fig. S24**). The data from Yale University has two 100X bulk samples and eight 30X fractions per brain. We performed five calling with different combinations of samples combined. We conducted more straightforward calling strategies for the other two data sets, either for a single 250X bulk sample or one combined of two 100X bulk samples from two brain regions.

Furthermore, all mutations were divided into tier1 and tier2. Tier1 had high confidence mutations called from bulk brain samples with the combined coverage of at least 200X. Tier2 had other mutations which were for brains of shallower coverage, of lower confidence, or made for samples representing sorted cell fractions. Most of the analyses were conducted using tier1 mutations, but some, as specified below, included tier2 mutations.

For the same set of brains sequenced at Harvard University, the generated call set (tier1+2) was 66.2% larger than the one reported previously (*6*) (**Fig. S1A**). At the same time, we re-discovered 63.7% of all and 85.2% of validated mutations reported previously (*6*), suggesting more sensitive yet accurate mutation call set.

#### Mutation phasing to haplotypes (contributed by Taejeong Bae)

Read-backed haplotype phasing information for mosaic candidate sites of each sample (**Fig. 1B**) was extracted using Phasing.py, a python script of MosaicForecast (*53*) with the following parameters.

~~~
python MosaicForecast/Phase.py \
      sample_bam_dir out_dir \
      resources/hs37d5.fa input.bed 20 \
      resources/hg19/k24.umap.wg.bw
~~~

#### Mutation signature analysis (contributed by Taejeong Bae)

We used a custom python script to draw mutational spectra for 6 mutation types of single base context (**Fig. 1C**) and for 96 mutation types of trinucleotide context (**Fig 1D, S1E**). Cosine similarity between two mutational spectra was calculated using the SciPy python library.

#### Functional annotation (contributed by Taejeong Bae)

To annotate functional consequences of discovered somatic mutations (**Tables 1&2, Fig. 4A**), we ran command line of Variant Effect Predictor (VEP) version 101.0 (*54*) with the following parameters.

~~~
vep --fork 12 --offline -assembly GRCh37 --format ensembl \
    --pick --pick_order rank,canonical,biotype,ccds,length \
    --sift b --polyphen b --ccds --symbol --numbers --regulatory
    --canonical --biotype --gene_phenotype \
    --force --tab -i input_call_set.txt -o variant_effect_output.txt \
    --stats_text -sf variant_effect_stats.txt
~~~

We considered missense, stop gain or loss, and splice related mutations as putative damaging mutations in our analyses. Out of two mutations in PCDH15, one in UK20119 (tier 2 mutations) was not validated by amplicon-seq and was not utilized for downstream analyses.

#### External datasets (contributed by Taejeong Bae)

To annotate cancer driver genes (**Table 1**), we used 723 genes from the COSMIC Cancer Gene Census v94 (*12*) and 299 genes from Bailey et al. (*11*). To annotate genes relevant to neuropsychiatric diseases (**Table 1**), we used several external data sources. For the known human genes associated with ASD, we used the SFARI Gene database (https://gene.sfari.org, release of 06-10-2021). For TS, we compiled genes from two previous studies (*26, 55*). Wang et al. (*26*) did a whole exome study providing genes with *de novo* mutations in TS patients. We selected 205 genes from Table S7 in that study with the criteria that dn.mis3 + dn.lof >= 1 and p-value < 0.05. Yu et al. (*55*) conducted a GWAS meta-analysis reporting 39 LD-independent index SNPs with p-values < 10^−5^. We extracted 187 genes associated with those index SNPs from Table S4 in that study. For SCZ, we collected SCZ-relevant genes from four previous studies (*13–16*). From whole exome studies, we gathered 25 genes with p-value < 0.05 from Tables 1&2 in Rees et al. (*13*) and 166 genes with p-values < 0.05 from Supplementary Data 11&12 in Howrigan et al. (*14*). From previous GWAS, we compiled 42 genes from Tables S12&S13 in Pardinas et al. (*15*) and 153 genes from Supplementary Table 4 in the study of the Schizophrenia Working Group of the Psychiatric Genomics Consortium (*16*).

#### Genotyping of mutations in single nuclei (contributed by Liana Fasching and Taejeong Bae)

About 0.2 g of striatal tissue (caudate nucleus and putamen) was dissected from the brain NC7 and nuclei were isolated as described above in “Isolation of nuclei and fractionation of bulk tissue”. Nuclei were stained against CTIP2 and NeuN and FAN-sorted. Single NeuN+/CTIP2-nuclei (corresponding to the STR-INT fraction) were collected into PCR strip tubes. For 16 single nuclei, Primary Template-directed Amplification (PTA) amplification was performed using the ResolveDNA^™^ Whole Genome Amplification Kit (Bioskryb Genomics). Amplified DNA was sequenced at 5X coverage. Sequencing reads were processed as described above. We considered the sites with two or more allelic read support as genotyped. Genotyping sensitivity was estimated as the genotype rate for germline heterozygous SNP with the same genotyping criteria requiring 2 or more supporting reads. Heterozygous SNPs were discovered using GATK by combining bulk data for cortex and striatum.

#### Comparing lineage distributions between brain regions (contributed by Taejeong Bae)

For each brain processed at Yale University, we generated sequencing data of bulk tissue from cortex and striatum and four cell fractions from each of the two brain regions. To assess an uneven distribution of lineages between cortex and striatum, we simulated the measured difference of mutation frequencies assuming that the mutations are distributed evenly, i.e., have the same VAF. Specifically, for each mutation, we had read coverage observed in each brain region and average VAF across all data for the brain as input parameters. We first simulated the coverage of a mutation in each region (by Poisson distribution around coverage from the data) and then simulated the count of alternative alleles for the average VAF (by Poisson distribution around expected allele counts, which is a product of simulated coverage by average VAF). We only retained outcomes with at least 5 alternative alleles to apply the same minimum supporting read count requirement used in the call filtering step. We repeated the simulation 10,000 times per brain to produce the simulated distribution of VAF differences. We then compared simulated and observed distribution of differences by Kolmogorov–Smirnov. With the same approach, we assessed an uneven distribution of lineages between cortex and hippocampus for the brains processed at LIBD. We found a significant bias of VAFs toward one tissue in 8 brains for the Yale cohort and 9 brains for the LIBD cohort (**Fig. S3B,E**).

For the 8 brains with biased VAFs in the Yale cohort, higher VAFs were consistently observed in the cortex except for the hypermutable brain NC7. Given difficulties isolating interneurons fraction from the cortex, analysis in NC7 could be biased because of the high frequency of mutations in striatum interneuron fraction in this brain. When excluding this fraction from calculations, higher VAFs of NC7 inclined toward the cortex (**Fig. S3C**). In the LIBD cohort, all 5 non-hypermutable brains among the 9 brains with biased VAFs showed higher VAFs leaned toward the cortex.

For the all non-hypermutable brains with biased VAF from the two cohorts (6+5), we tested the significance of the enrichment of higher VAFs toward cortex by binomial test assuming the same chance of bias toward cortex or non-cortex tissue (p-value = 5×10^−4^).

#### Deriving a set of developmental brain enhancers from a telencephalic organoid model (contributed by Scott Norton)

To understand the epigenetic landscape of brain organoids in ASD, we generated telencephalic organoids form 11 pairs of ASD probands and their unaffected fathers expanding our previous dataset (*48*). DNA was collected to map histones H3K27ac, H3K27me3, H3K4me1, and H3K4me3 via ChIP-seq at 0, 30, and 60 days following induction of terminal differentiation (TD) by withdrawal of growth factors from the media. All four ChIP-seq marks were used to train a nine-state segmentation model in ChromHMM (*56*), and states were annotated according to their emission probabilities using the Epigenome ROADMAP 18-state model as a reference (*57*). H3K27ac ChIP peaks from 54 experiments were called using MACS2, and consensus regions were defined as those with at least 1 bp overlap in at least three original (per-library) peaks. This resulted in 654,738 H3K27ac consensus peaks, of which 436,296 have majority overlap with enhancer states (bivalent, weak, active1, active2) in the segmentation. Enhancers were linked to gene promoters either by proximity on the linear genome or by inferred gene-enhancer loops from published Hi-C datasets from fetal brain and/or human cell lines as described previously (*48*), resulting in a curated set of 204,220 gene-linked enhancers. These enhancers were further filtered by groupwise H3K27ac activity and expression of the putative target gene, then lifted over to hg19 for a final list of 200,000 high-confidence gene-linked enhancers (**Table S6)**.

#### Analyzing mutation enrichment in enhancers (contributed by Yifan Wang)

We overlapped somatic point mutations identified from all samples excluding the ones from hypermutable brains with the gene-linked enhancers from brain organoids (see below), from developing human brain (*58*), and from induced neurons (*59*). We then tested the enrichment of somatic point mutations from different diseases comparing to controls using Fisher Exact test. Next, we tested the possible function effect of the somatic point mutations within gene-related enhancers regions by analyzing the disruption of transcription factor binding motifs by the mutations. We detected all motifs disrupted by the mutations with a position weight matrix (PWM) similarity score (equation see below) larger than 0.7 (*60*). We then computed the score change of the somatic point mutations (equation see below). We preserve all the disruption of somatic mutations with a score change larger than 0.5 for the downstream analysis.

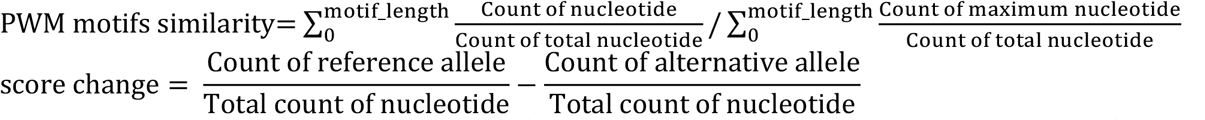

We then compared the number of motifs disrupted by somatic point mutations from TS, ASD and SCZ samples to control samples with a binomial test taking the total number of somatic point mutations into consideration (**Fig. 4B**).

#### Method for calling large mutations (contributed by Milovan Suvakov)

To detect large CNVs we used CNVpytor (https://www.biorxiv.org/content/10.1101/2021.01.27.428472v1). CNVpytor imports information about read-depth (RD) from alignment file and information about B-allele frequency BAF based on SNPs from variant (VCF) file. Two-dimensional likelihood function within large bins (e.g., 10 kbp or 100 kbp) is calculated as product of two independent functions based on RD and BAF. For RD likelihood we used gaussian function centered around mean RD value with calculated standard deviation that includes local and global fluctuations. BAF likelihood is calculated as product of symmetrized beta distributions for each SNP. Segmentation of genomic regions with similar RD and BAF is calculated by merging neighboring bins with maximal overlap between two-dimensional likelihood functions. After merging, composite likelihood function is assigned to new segments. Iterations end when there are no additional significant overlaps between two neighboring segments (multiple hypothesis p-val < 0.05). During next phase CNVpytor continues merging not only neighboring segments but segments that are close enough (gap between them is smaller than 1/3 of total length). The tool outputs a list of all significant segments with p-values based on RD and BAF, most likely copy number model (i.e., copy number for each haplotype), and corresponding cell frequency. When cell frequency and size of the event are rather small (cell frequency below 20% and size less or about 100 kbp), multiple copy number models can have a similar likelihood, and CNN-LOH can be confused with deletion or duplication. Additional evidence, such as discordant read pairs for deletions and duplication can confidently resolve event type in such cases.

We ran CNVpytor 1.1 with the following commands:

~~~
cnvpytor -root sample.pytor -log sample.log -j 24 -rd sample.cram -T hs37d5.fa -chrom $(seq 1 22) X Y MT
cnvpytor -root sample.pytor -log sample.log -his 500 1000 10000 100000 1000000
cnvpytor -root sample.pytor -log sample.log -snp sample.ploidy_2.vcf.gz
cnvpytor -root sample.pytor -mask_snps
cnvpytor -root sample.pytor -baf 10000 100000
cnvpytor -root sample.pytor -call combined 100000 > sample.2d.100000.txt
cnvpytor -root sample.pytor -call combined 10000 > sample.2d.10000.txt
~~~

For samples from male brains, we evaluated copy number of X chromosomes by running standard read depth segmentation algorithm using bin size of 10000 bp. The command was as follows: cnvpytor -root sample.pytor -call mosaic 10000 -chrom X -rd_use_mask

For samples from male brains, we evaluated copy number of Y chromosomes by comparing the read depth of with that of a region on X chromosomes using bin size of 1000 bp. The command was as follows:

~~~
echo “compare Y X:25M-35M” | cnvpytor -root sample.pytor -view 1000 -rd_use_mask
~~~

#### Calling large mutations (contributed by Yifan Wang)

UMB5308 from the ASD cohort was excluded from the analysis due to the high noise of the sample. LIBD82, a brain with a potential tumor (**Fig. 1E**), was excluded from the CNV analysis. Other samples with a combined coverage less than 100X were excluded from the analysis, as having relatively shallow coverage to discover large mutations. In total, we performed the analysis on 59 ASD brains, 9 SCZ brains, 19 TS brains, and 44 normal brains. Calls for large mutations were generated by CNVpytor (https://www.biorxiv.org/content/10.1101/2021.01.27.428472v1) with the caller gathering information from both read depth and split in B-allele frequency of germline SNPs. SNPs were called using GATK haplotype caller run with ploidy=2. Analysis was conducted with two bin sizes: 100 kbps and 10 kbps.

We then applied filters to exclude false positives and germline CNVs. We considered as false positives the following: 1) calls with adjusted p-value from CNVpytor larger than 0.05/(number of samples*3*10^9^/bin size); 2) calls with <50% of well mapped bases (P-bases) as defined by the 1000 Genomes Project; 3) calls with >5% of non-sequenced reference (N-bases); 4) calls only supported by read depth (p-value from BAF signal > 0.01) and with predicted cell frequency <5%; 5) calls with predicted cell frequency <10%; 6) calls found in multiple samples (two calls are considered the same if overlap by 50% reciprocally). We additionally filtered out calls with length <= 3 of bins due to the boundary effects, which may lead to underestimation of cell frequency for germline CNVs. We, then, manually filtered out 16 germline CNVs in complex genomic regions (**Fig. S19**).

For the identified large mutations, we applied Manta (*61*) to resolve breakpoints. We ran Manta 1.6.0 with a random sample within each cohort as the control within the region 3 times larger than the mutations size (1 size upstream and 1 size downstream). The command to ran Manta was as follows:

~~~
manta-1.6.0-0/bin/configManta.py \
      --normalBam control.cram \
      --tumorBam sample.cram \
      --referenceFasta hs37d5.fa \
      --runDir sample.dir \
      --callRegions region.bed.gz
~~~

For the somatic large mutations with no resolved breakpoints, we imputed two haplotypes by phasing germlines SNPs using population haplotypes (see **below**) and then confirmed that the frequencies of the two haplotypes were different (**Table S4, Figs. S5-S18**).

#### Phasing of germline SNPs to haplotypes (contributed by Taejeong Bae)

For each individual, the phasing was conducted using shapeit4 (version 4.2.0, https://odelaneau.github.io/shapeit4) with the following command and parameters

~~~
shapeit4 -T 20 \
     -I unphased.${chr}.vcf \
     -H ALL.chr${chr}.phase3_shapeit2_mvncall_integrated_v5a.20130502.genotypes.vcf.gz \
     -M chr${chr}.b37.gmap.gz \
     -R ${chr} \
     -O phased.${chr}.vcf
~~~

The genetic maps for humans were downloaded from https://github.com/odelaneau/shapeit4/tree/master/maps. For the reference panel data, we used 1000 Genomes Phase 3 haplotypes obtained from ftp://ftp.1000genomes.ebi.ac.uk/vol1/ftp/release/20130502.

#### Validation of mutations in samples from Yale University (contributed by Liana Fasching, Taejeong Bae, and Yifan Wang)

Several mutations discovered in TS and NC brains were validated by amplicon-seq in the following samples: 4 point mutations in TS9 in STR-bulk and in CX-bulk (BA6, BA9 lateral, BA9 medial, BA17, BA44/45 and SMA) and 2 point mutations in NC7 in STR-bulk and in CX-bulk (BA6 and BA9). Duplications were validated in NC6 (STR-bulk and BA6 CX-bulk) and in TS1 (STR-bulk and BA6 and BA17 of CX-bulk). All mutations were tested and invalidated in control DNA from cell line NA12878.

We determined the genomic location of each point mutations using the 1000 Genome Browser (GRCh37.p13/hg19 https://www.ncbi.nlm.nih.gov/variation/tools/1000genomes/). Primer pairs were designed using Primer3 (http://primer3.ut.ee). Specificity of primers was confirmed by UCSC *in silico* PCR (http://genome.ucsc.edu/cgi-bin/hgPcr). <u>Primer pairs for duplications were designed around their break points using the same method. All primers are listed in</u> (**<u>Table S7</u>**).

Phusion High-Fidelity DNA polymerase (ThermoFisher) was used for PCR amplifications. Size and presence of each amplicon was confirmed by gel electrophoresis (2% agarose gel). Amplified DNA fragments were purified using the QIAquick PCR Purification Kit (Qiagen). Samples were multiplexed and sequenced under following conditions: MiSeq, 2×300 bp reads.

Sequencing reads were aligned to the human reference genome GRCh37d5 (ftp://ftp-trace.ncbi.nih.gov/1000genomes/ftp/technical/reference/phase2_reference_assembly_sequence/hs37d5.fa.gz) using bwa (version 0.7.17) and sorted with sambamba (version 0.6.7). For validation of point mutations, the variant allele frequency and allelic count of each site were obtained using a custom python script with base quality 20 and mapping quality 20 and compared to ones obtained from the control sample NA12878. We further required coverage of at least 150,000X per site. Out of all mutations tested the lowest frequency in all brain samples was 0.7%, while in control DNA (NA12878) it was never above 0.02%. Based on these we consider all mutations as validated.

Breakpoints for duplications identified from TS1 and NC6 were reconstructed by Manta (*61*). Paired-end sequencing reads were then aligned to the reconstructed breakpoint sequence in addition to hg19 reference genome. Reads mapped to the breakpoint with a mapping quality larger than 30 were counted as supportive evidence for somatic duplications. Both duplications were validated based on comparison with read support from control DNA (NA12878) as depicted in **Figs. 4** and **S5**.

#### Validation of mutations by single cell sequencing (contributed by Taejeong Bae)

In brain NC7, nearly all the discovered mutations were genotyped (after adjusting for sensitivity) in the genomes of 8 nuclei extracted from the striatal STR-INT fraction. Single nuclei were amplified by the PTA approach and sequenced at 5X as described in the section “Genotyping of mutations in single nuclei”. This observation proved that the mutations belonged to the same cell lineage represented by those 8 cells. Based on this premise, we combined sequencing data from the 8 single nuclei to achieve combined whole genome coverage of over 40X for that lineage. We considered mutations as validated if their VAF in the data was at least 30%. With this criteria, 833 mutations (323 tier1 mutations) were validated in this lineage. These validated mutations include all the four putatively damaging mutations in genes previously implicated in cancers (**Table 1**).

#### Validation of mutations in samples from Harvard University (contributed by Caroline Dias)

Mutations were validated with the Genewiz Amplicon EZ protocol as follows. For *PCDH15* mutation in case UMB4231 and *MTOR* mutation in case AN05983, primer pairs were designed with Primer3 to flank the region of interest and specificity verified with in silico PCR as above. Illumina adaptors were added as per the Genewiz Amplicon-EZ specifications, and all primer pairs were tested with control genomic DNA to ensure a single band was generated in PCR as expected. Approximately 25 mg of frozen brain tissue was dissected into 3-4 pieces on a -20 cryostat, and genomic DNA extracted with the DNEasy Tissue kit. Following Phusion High Fidelity PCR, the PCR product was confirmed with gel electrophoresis and purified with the Purelink PCR clean-up kit. Picogreen was used to determine DNA concentration and samples were concentrated to 20 ng/ul. Following 2×250 bp sequencing and filtering (>66,000 reads for *PCDH15* amplicon and >177,000 reads for *MTOR*), mutations of interest were deemed validated when compared to background sequencing error rate.

**Figure S1.**
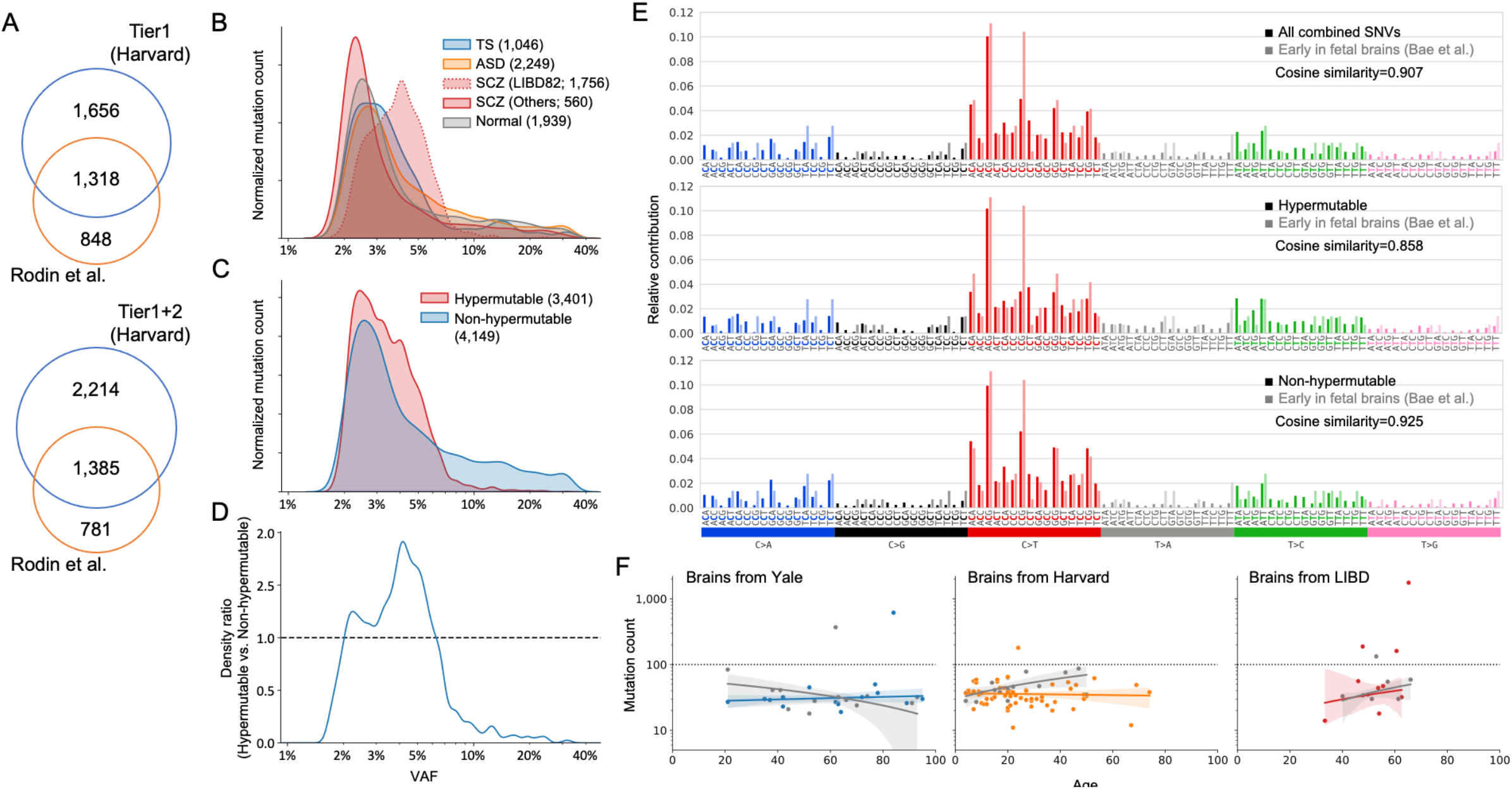
**A)** Overlap of somatic mutations discovered here and reported previously for brain sequenced at Harvard University (Rodin et al., Nature Neuroscience, 2021). We rediscovered 167 mutations (161 in tier1 and 6 mutations in tier2) out of 196 validated in Rodin et al. **B)** Distributions of somatic mutation by allele frequencies across cohorts, normalized to total counts of mutations. **C)** Normalized distributions of mutation allele frequencies according to hypermutable phenotypes. **D)** Ratio of mutations in hypermutable brains to those in non-hypermutable brains versus allele frequencies. **E)** Combined mutation spectrums for three sets of mutations: somatic mutations in all brains, mutations in hypermutable brains, and mutations in non-hypermutable brains. The spectrums are the same. **F)** Mutation burdens depending on the age of each brain.

**Figure S2.**
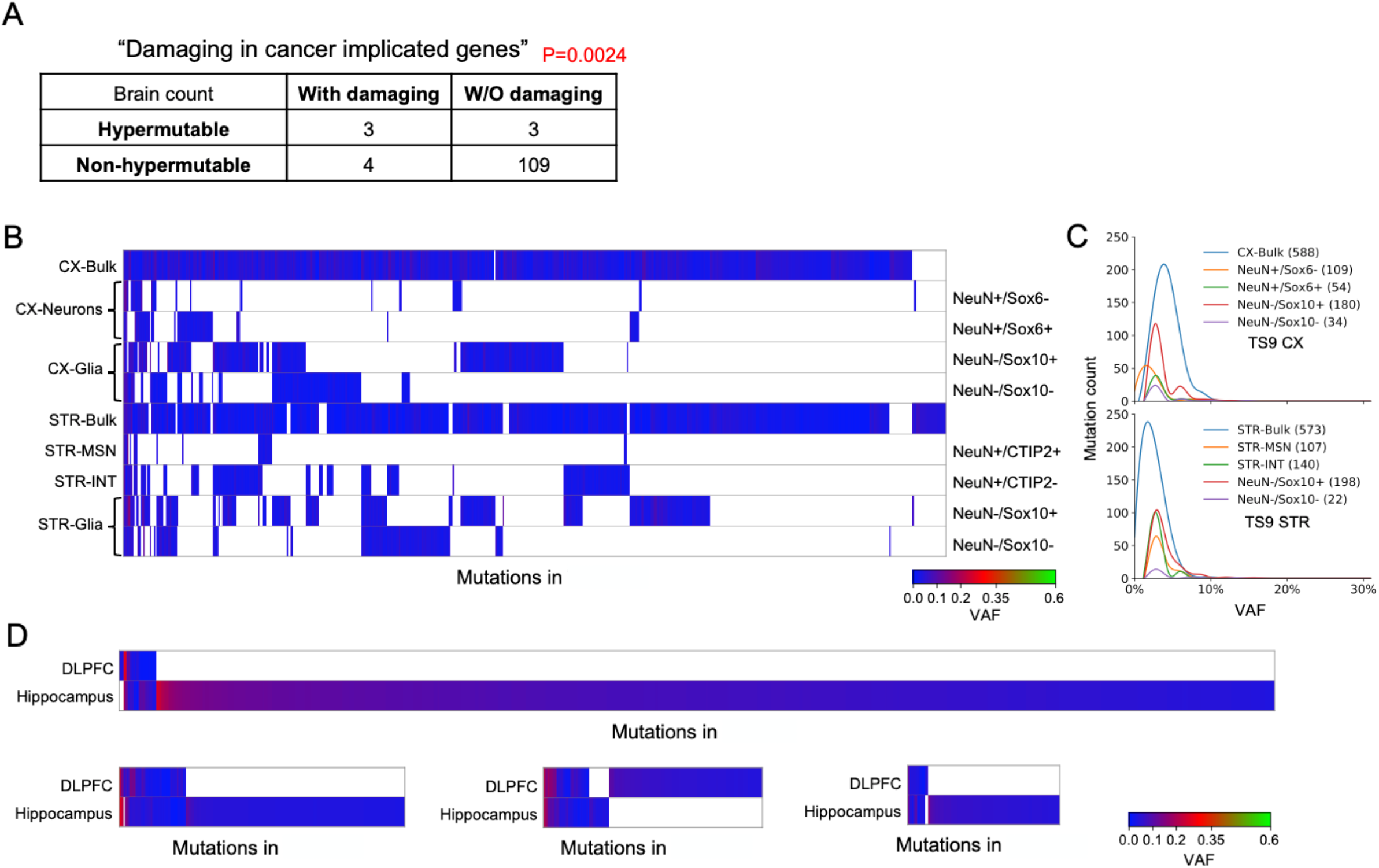
**A)** Contingency tables and p-values for Fisher’s exact test for enrichment of hypermutable brains with damaging mutations in cancer driving genes. **B)** Heat map of mutation allele frequency across samples in brain. **C)** Distribution of mutation allele frequencies across samples in brain. **D)** Heat map of mutation allele frequency across samples in brains.

**Figure S4.**
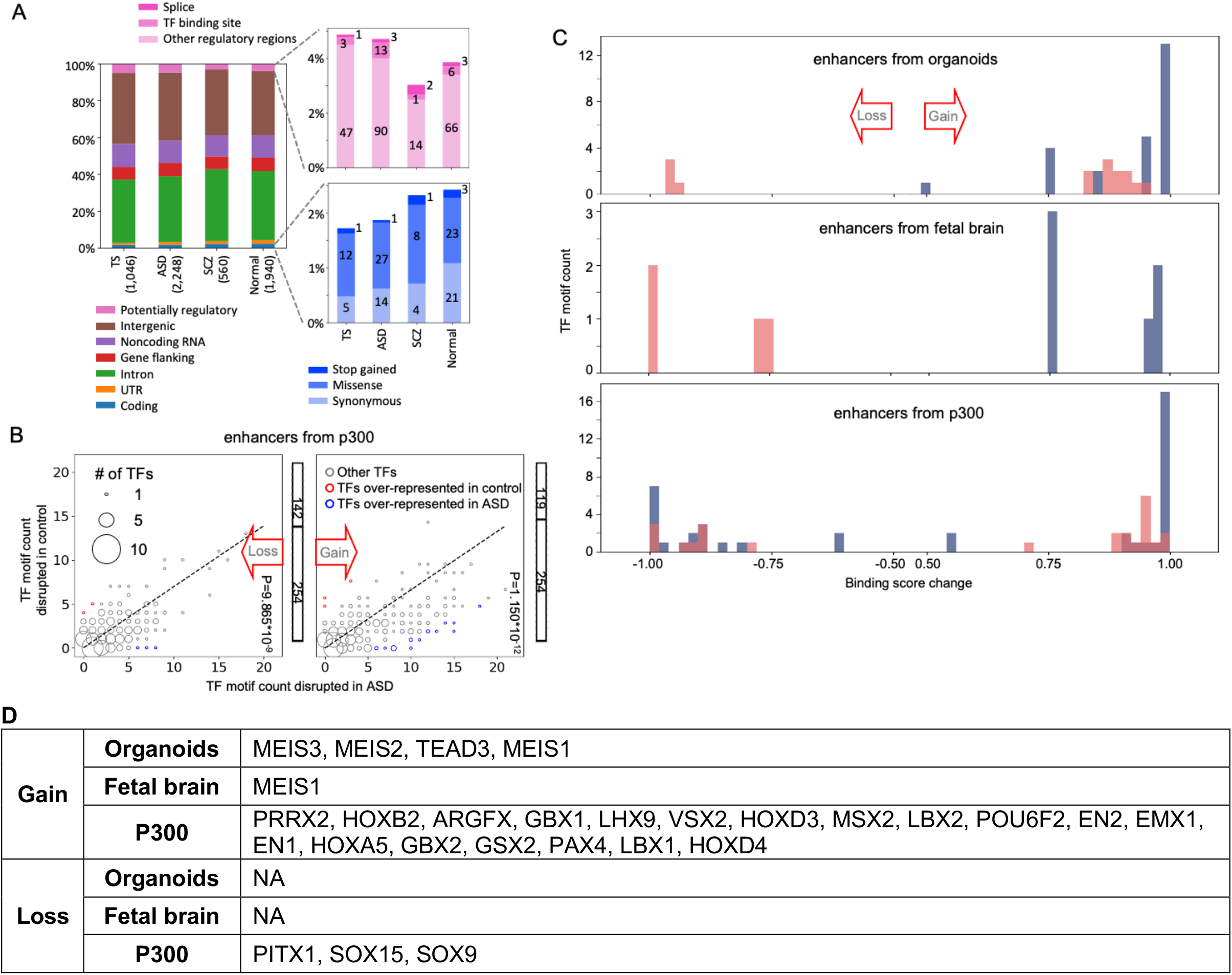
**A)** Functional impact category annotated by VEP including mutations in both hypermutable and non-hypermutable brains. **B)** Somatic point mutations within enhancers mapped by ENCODE using p300 ChIP-seq in induced neural cell lines {PMID: 22955616} and fetal brains {PMID: 23375746}. Mutations in ASD disrupts more TF binding sites. **C)** Difference in binding score for gain or loss of TF binding upon somatic point mutations in binding motifs of select TFs. TFs with significant differences in counts are colored (blue and red) in B. **D)** List of TFs with significant differences in counts (i.e., TFs colored blue and red in panel C).

**Figure S5.**
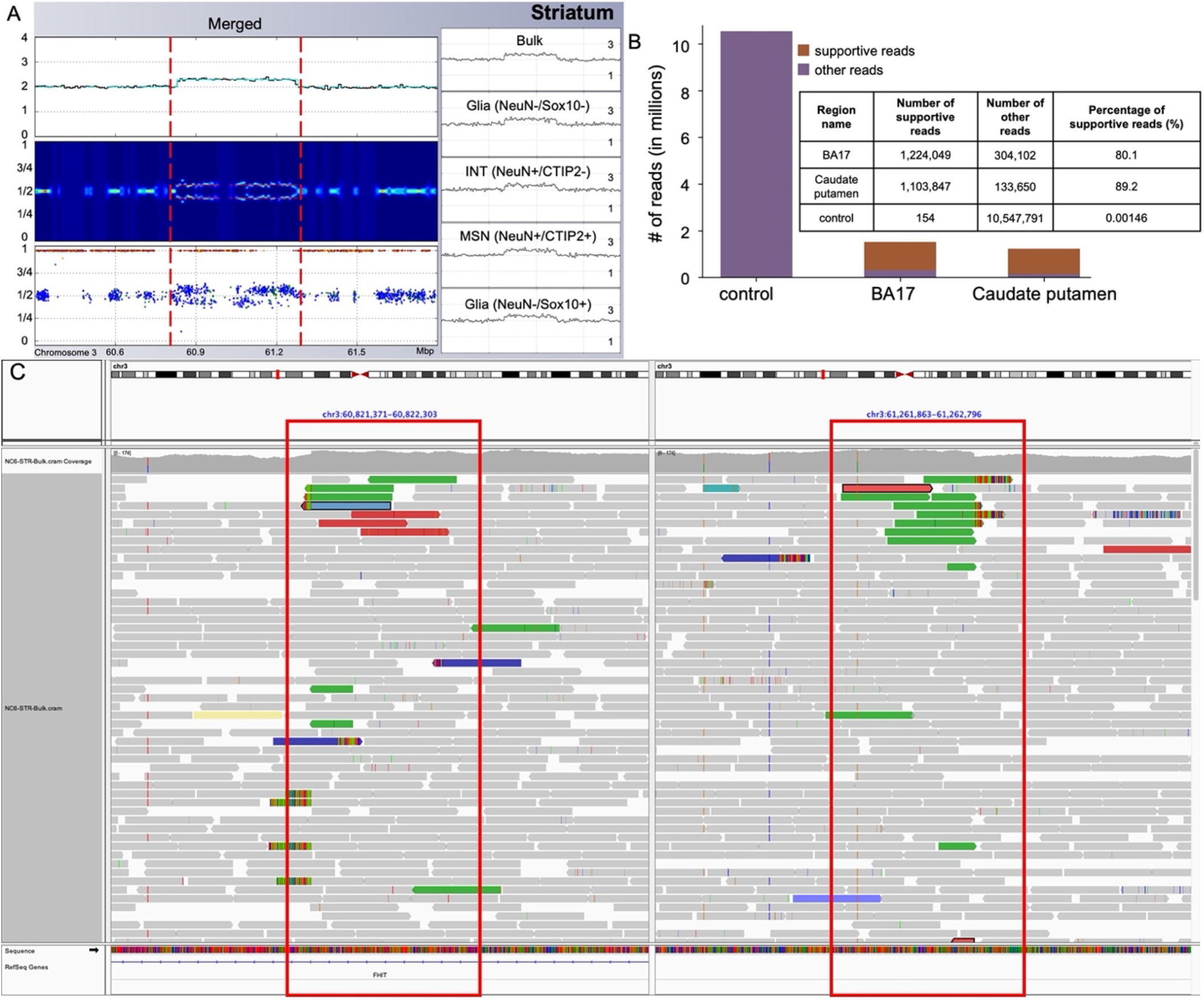
Somatic duplication on chromosome 3 (chr3:60822011-61262613) identified in. **A)** Increase of read depth, split of likelihood and allele frequency for individuals SNPs calculated from combined fractions and bulk data. **B)** Amplicon-seq validation of the duplication in A). Bar plot and table show the number of reads mapping to the junction of the duplication in the cortical region BA17 and caudate putamen of and in control sample. **C)** IGV screen shot of discordant read pairs (green reads) supporting the somatic duplication.

**Figure S6.**
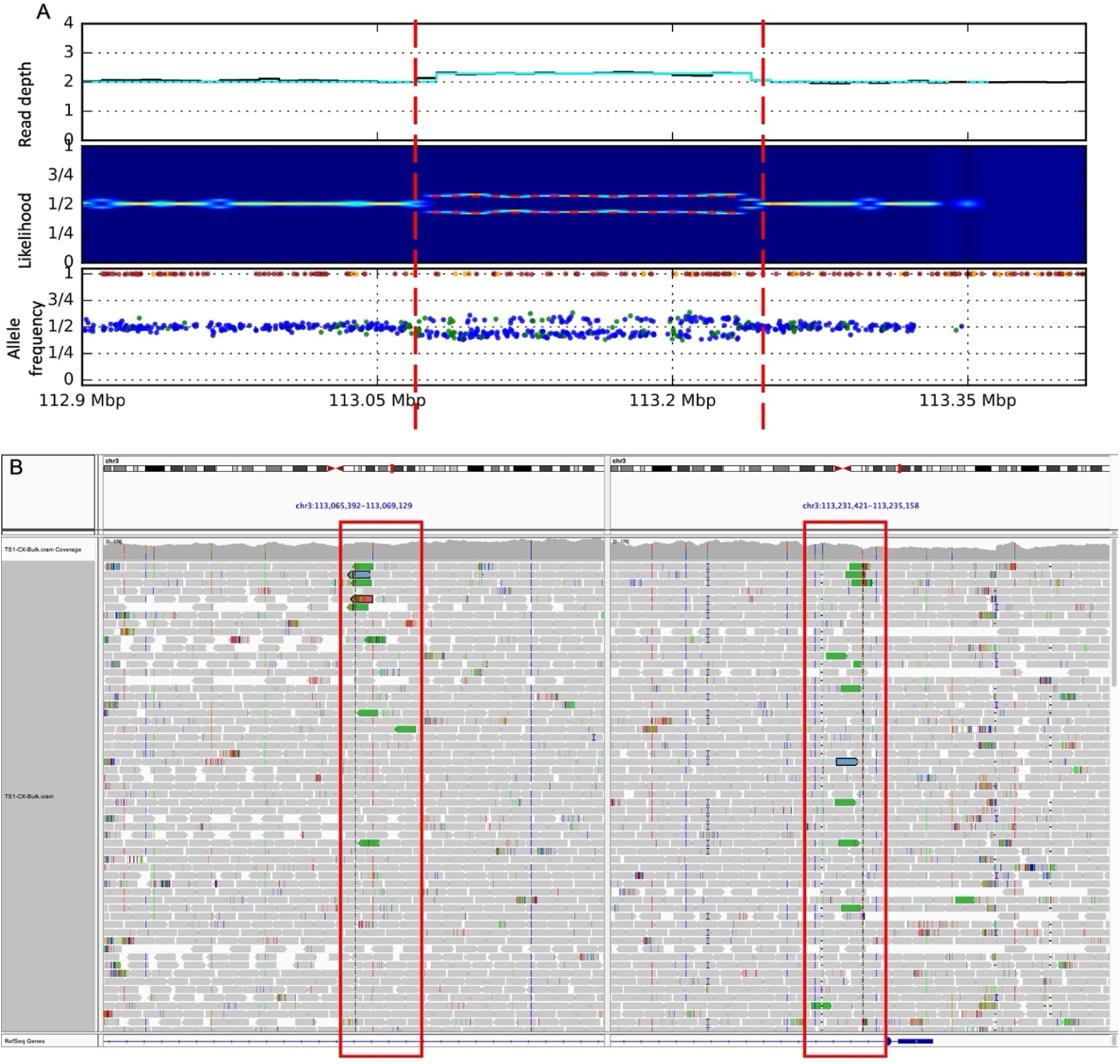
Somatic duplication on chromosome 3 (chr3:113067261-113233476) identified in. **A)** Increase of read depth, split of likelihood and allele frequency for individuals SNPs calculated from combined fractions and bulk data. **B)** IGV screen shot of discordant read pairs (green reads) supporting the somatic duplication.

**Figure S7.**
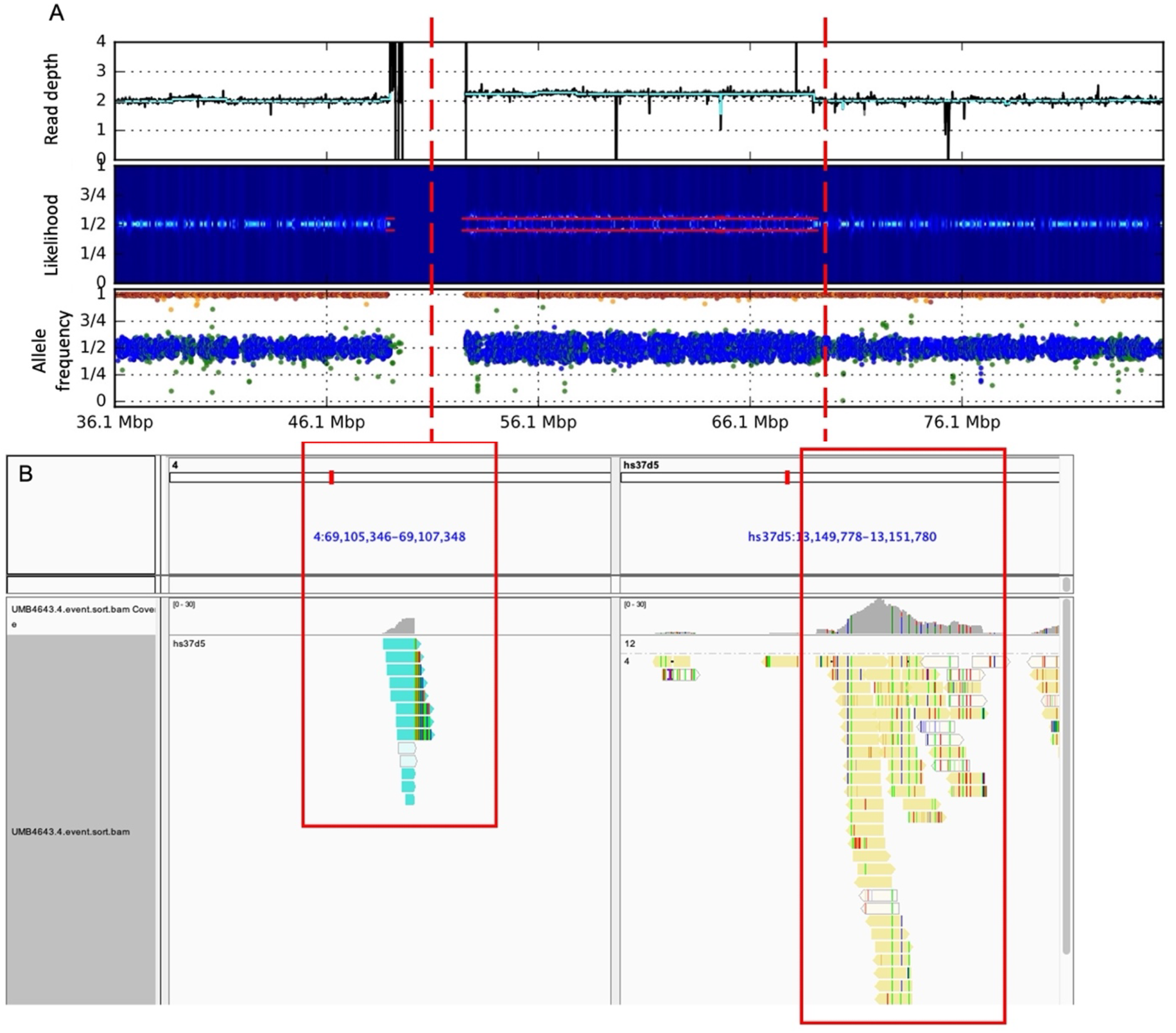
Somatic duplication on chromosome 4 (chr4:centromere-69106444) identified in. **A)** Increase of read depth, split of likelihood and allele frequency for individuals SNPs calculated from combined fractions and bulk data. **B)** IGV screen shot of discordant read pairs (turquoise blue reads on the left on chromosome 4 and lightyellow reads on hs37d5 contig) supporting the somatic duplication.

**Figure S8.**
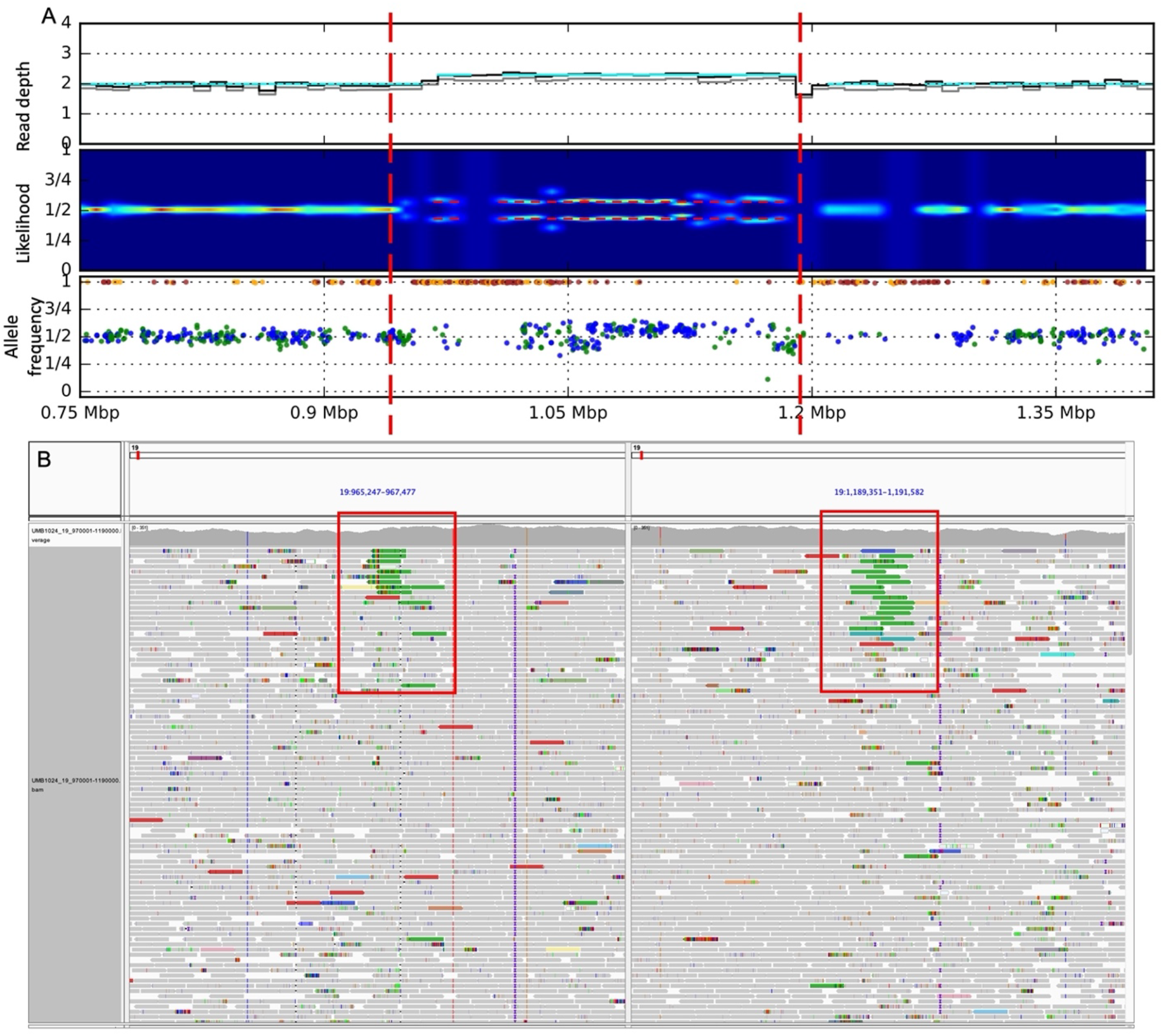
Somatic duplication on chromosome 19 (chr19:966363-1190674) identified in. **A)** Increase of read depth, split of likelihood and allele frequency for individuals SNPs calculated from bulk data. **B)** IGV screen shot of discordant read pairs (green reads) supporting the somatic duplication.

**Figure S9.**
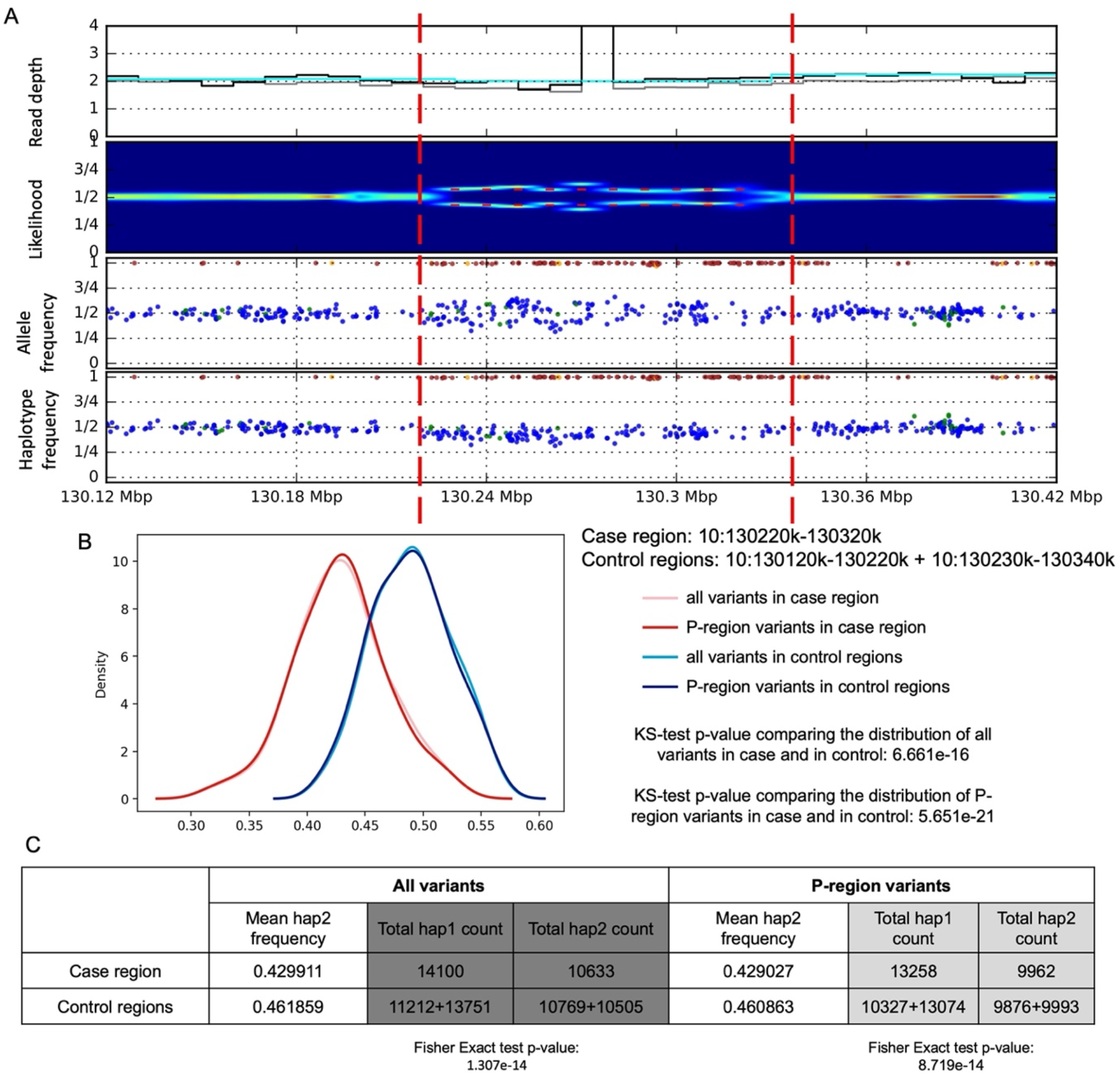
Somatic loss of heterozygosity (LOH) on chromosome 10 (chr10:130220001-130320000) identified in. **A)** Read depth, split of likelihood, allele frequency and one-sided haplotype phased allele frequency from combined data. **B)** Haplotype aware distribution of allele frequencies for SNPs within the predicted somatic LOH region (case, in red) and within upstream and downstream regions (control, in blue). The two distributions are different by KS-test (P-regions are defined by the 1000 Genome Project as accessible to short read sequencing). **C)** Table of read counts for two imputed haplotypes. The imbalance in case compared to control region is significant by Fisher Exact test.

**Figure S10.**
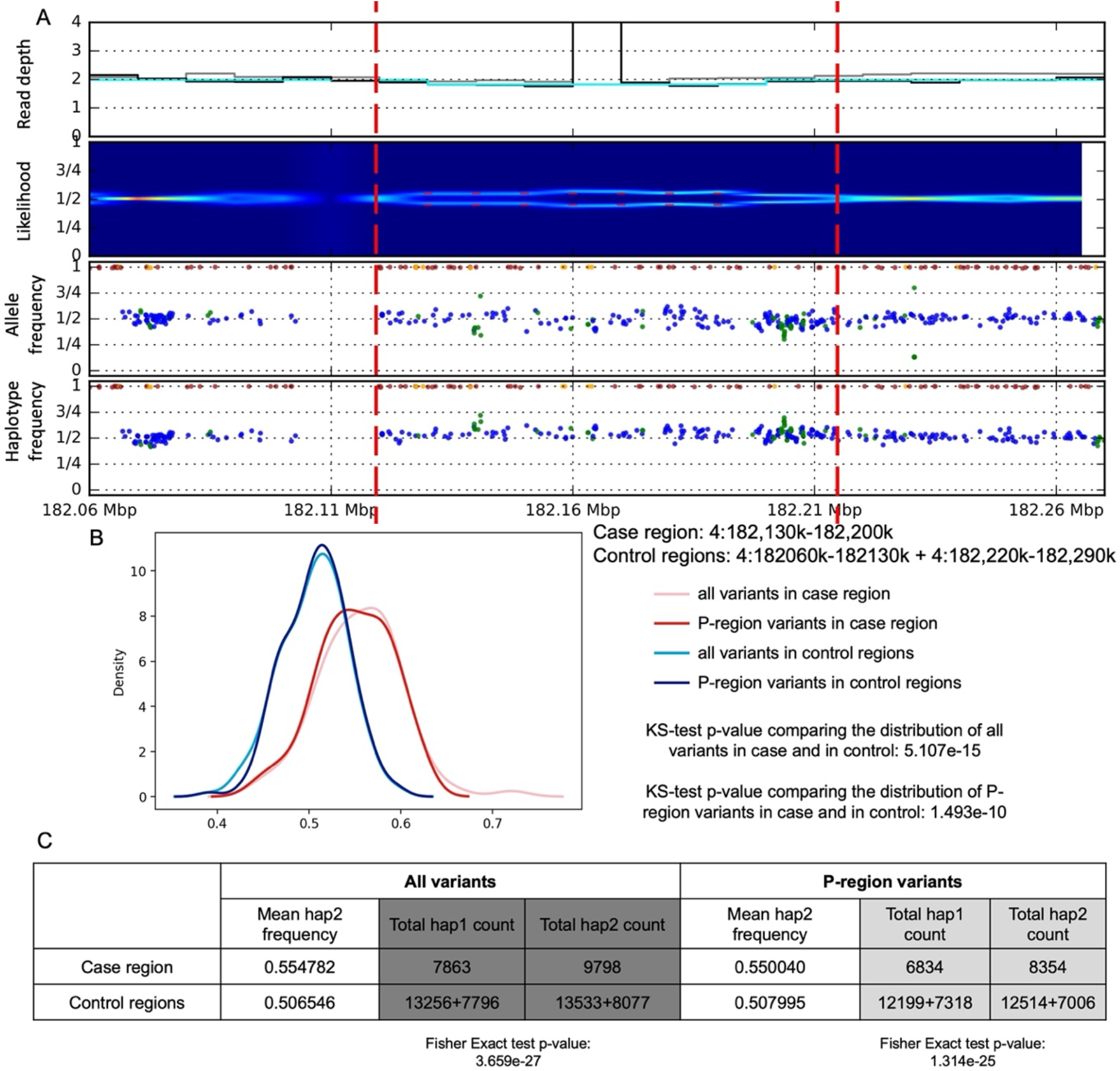
Somatic deletion on chromosome 4 (chr4:182130001-182200000) identified in. **A)** Read depth, split of likelihood, allele frequency and one-sided haplotype phased allele frequency from bulk data. **B)** Haplotype aware distribution of allele frequencies for SNPs within the predicted somatic deletion region (case; in red) and within upstream and downstream regions (control; in blue). The two distributions are different by KS-test (P-regions are defined by the 1000 Genome Project as accessible to short read sequencing). **C)** Table of read counts for two imputed haplotypes. The imbalance in case compared to control region is significant by Fisher’s Exact test.

**Figure S11.**
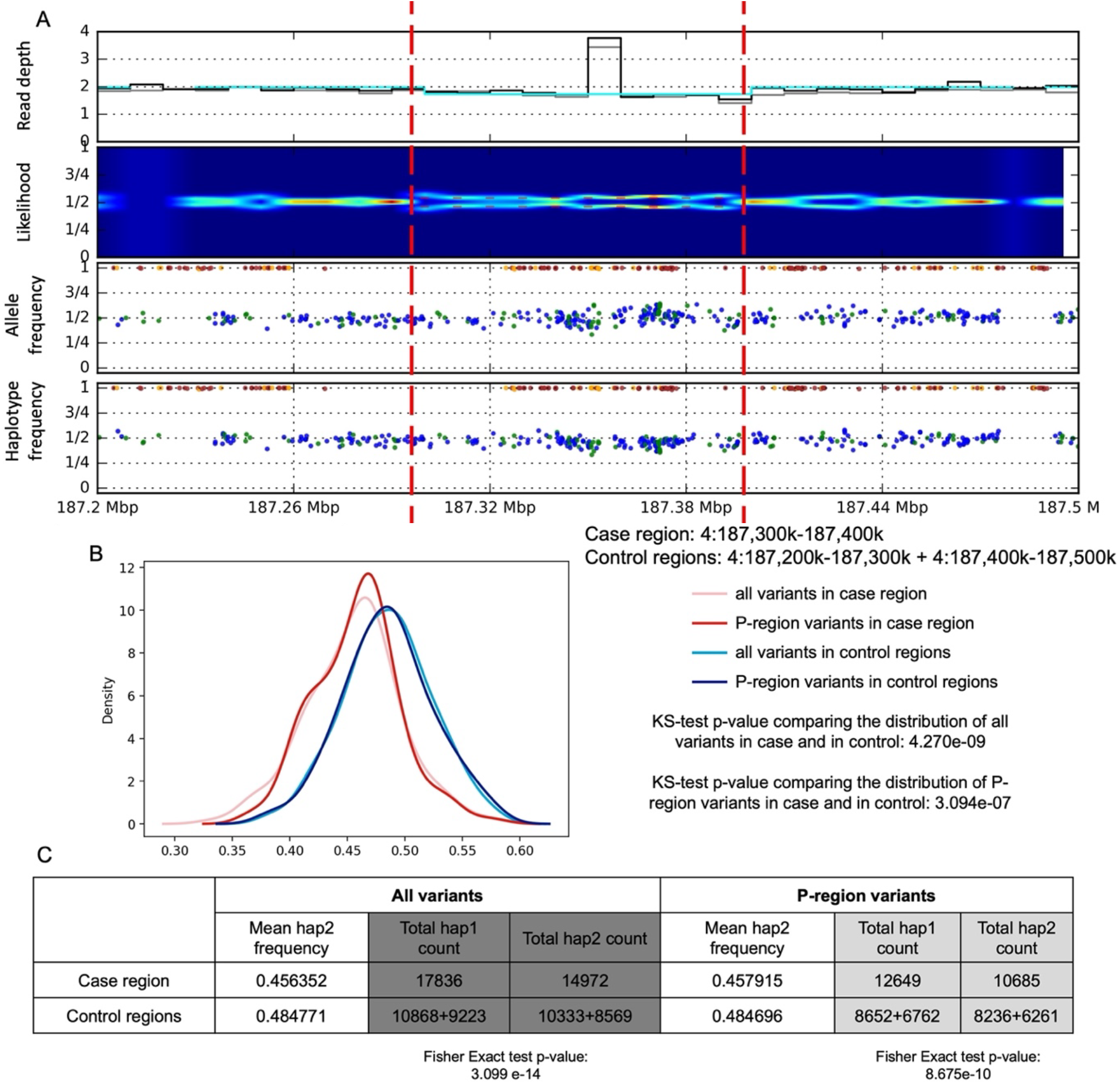
Somatic deletion on chromosome 4 (chr4:187300001-187400000) identified in. **A)** Read depth, split of likelihood, allele frequency and one-sided haplotype phased allele frequency from bulk data. **B)** Haplotype aware distribution of allele frequencies for SNPs within the predicted somatic deletion region (case; in red) and within upstream and downstream regions (control; in blue). The two distributions are different by KS-test (P-regions are defined by the 1000 Genome Project as accessible to short read sequencing). **C)** Table of read counts for two imputed haplotypes. The imbalance in case compared to control region is significant by Fisher’s Exact test.

**Figure S12.**
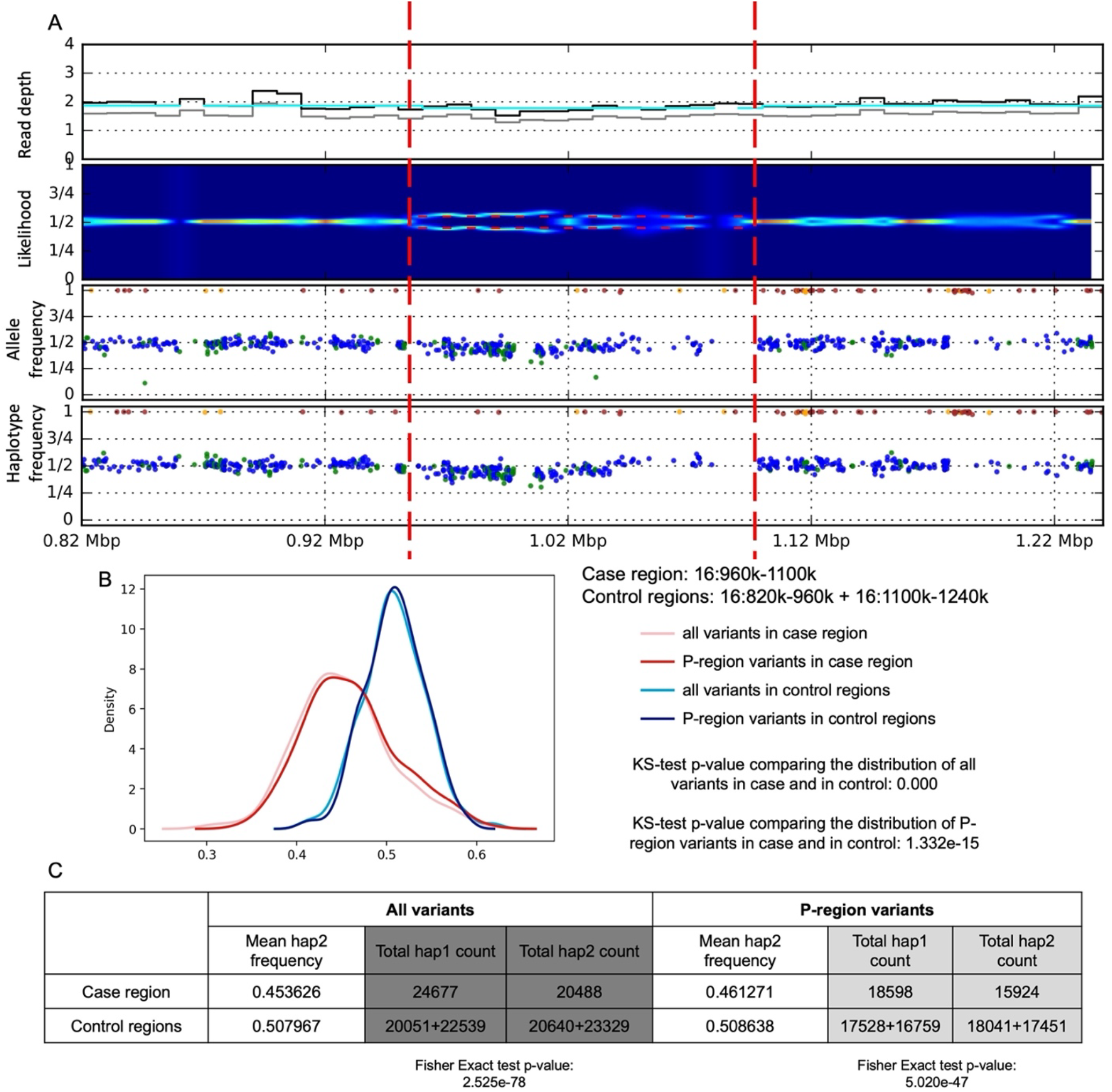
Somatic deletion on chromosome 16 (chr16:960001-1100000) identified in. **A)** Read depth, split of likelihood, allele frequency and one-sided haplotype phased allele frequency from bulk data. **B)** Haplotype aware distribution of allele frequencies for SNPs within the predicted somatic deletion region (case; in red) and within upstream and downstream regions (control; in blue). The two distributions are different by KS-test (P-regions are defined by the 1000 Genome Project as accessible to short read sequencing). **C)** Table of read counts for two imputed haplotypes. The imbalance in case compared to control region is significant by Fisher’s Exact test.

**Figure S13.**
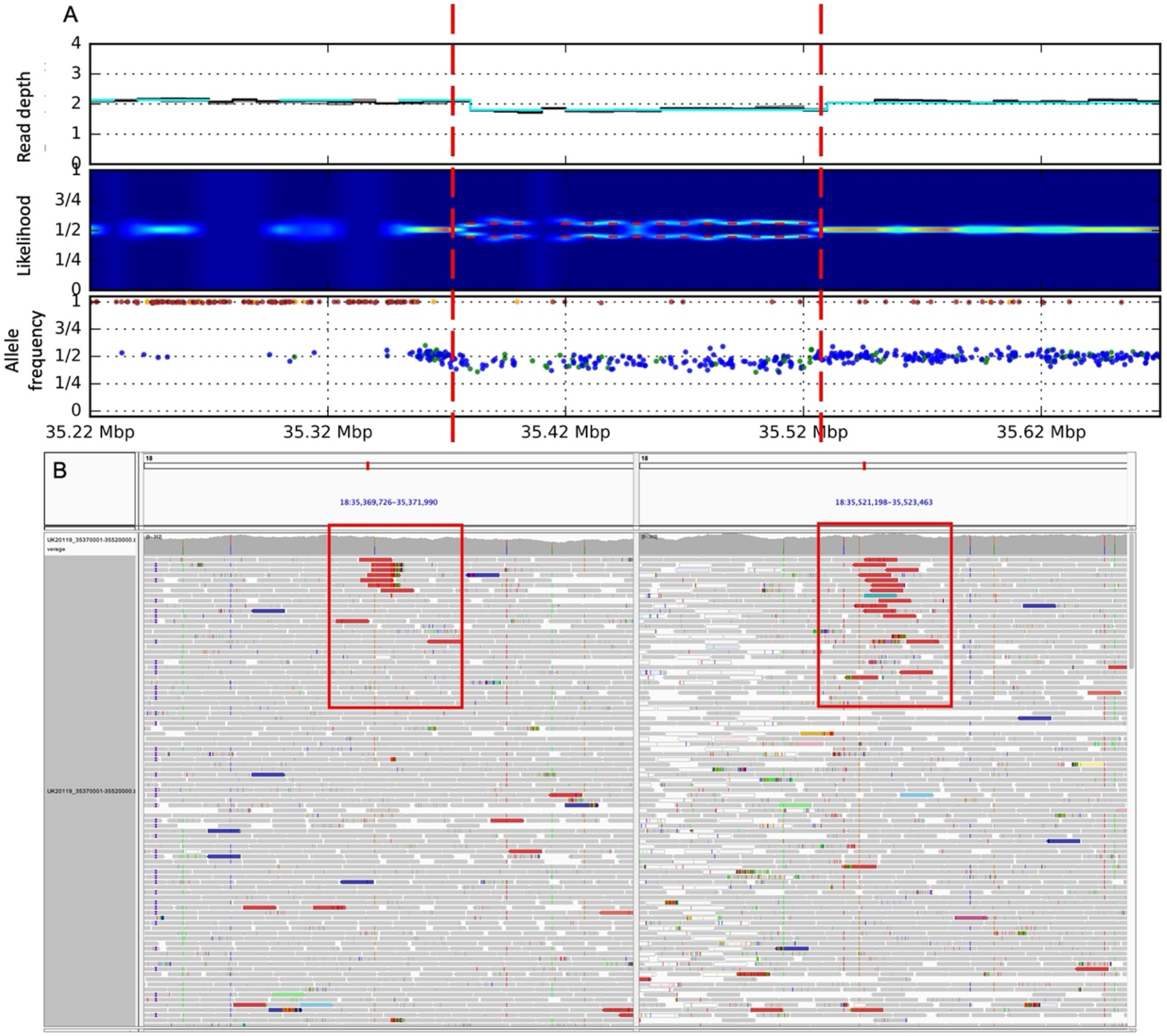
Somatic deletion on chromosome 18 (chr18:35370859-35522179) identified in. **A)** Increase of read depth, split of likelihood and allele frequency for individuals SNPs calculated from bulk data. **B)** IGV screen shot of discordant read pairs (green reads) supporting the somatic deletion.

**Figure S14.**
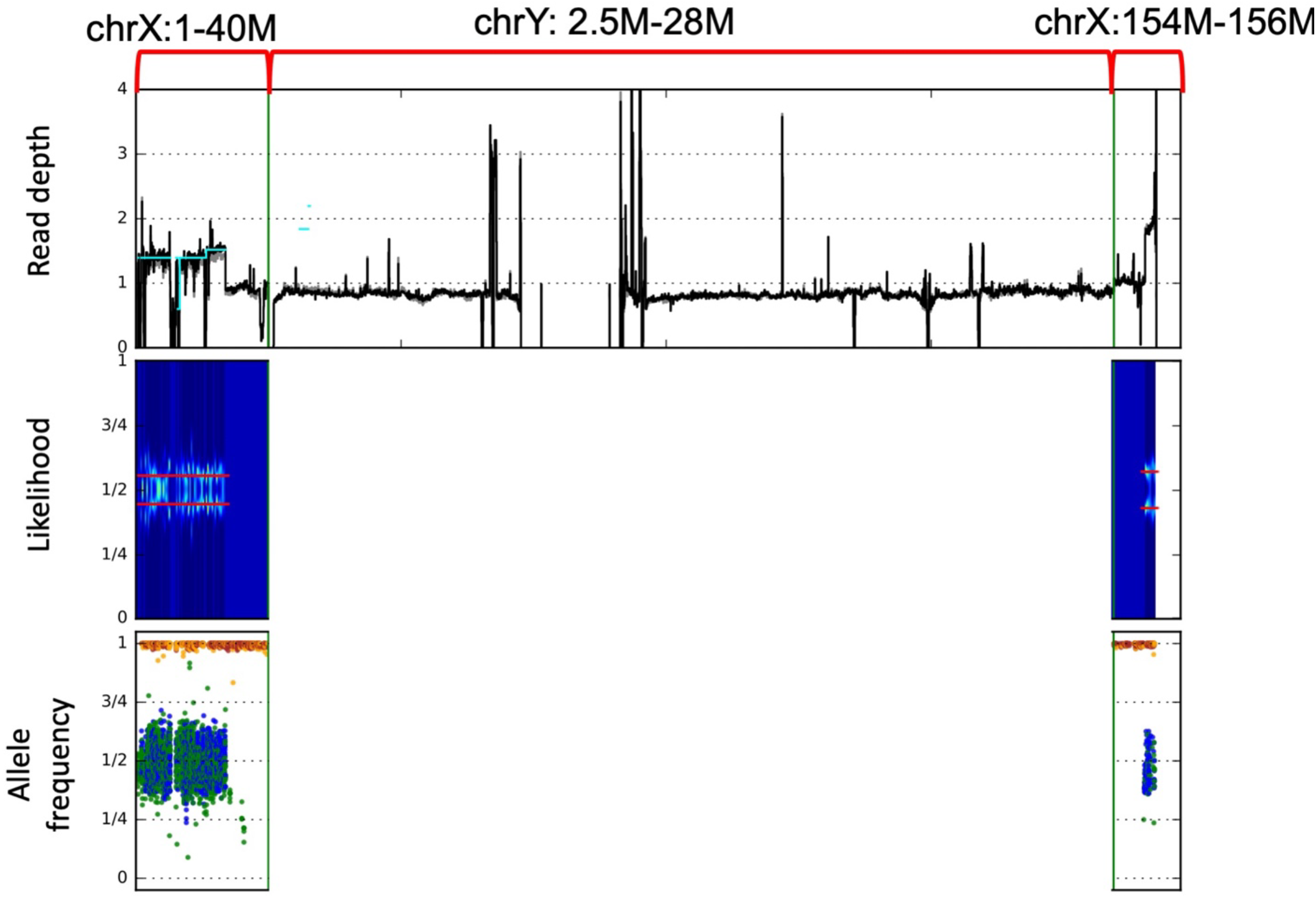
Somatic deletion of whole chromosome Y identified in. Decrease of read depth, split of likelihood and allele frequency within the pseudoautosomal region on chromosome X (PARI: chrX:1-27M), chrY (2.5M-28M), and pseudoautosomal region II on chromosome X (PARII: chrX:154-156M) from sequencing data of individual.

**Figure S15.**
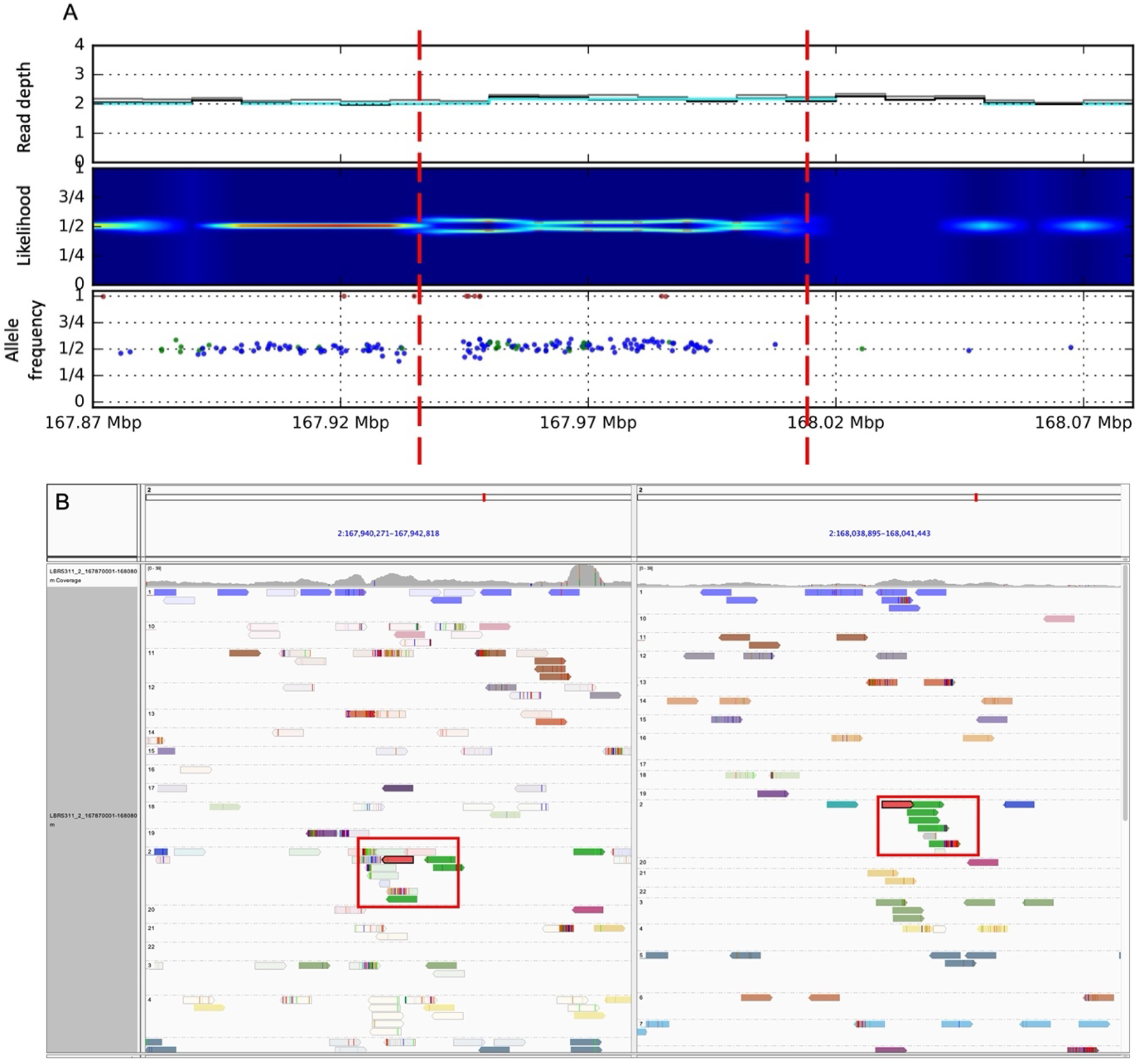
Somatic duplication on chromosome 2 (chr2:167940001-168010000) identified in. **A)** Increase of read depth, split of likelihood and allele frequency for individuals SNPs calculated from bulk data. **B)** IGV screen shot of discordant read pairs (green reads) supporting the somatic duplication.

**Figure S16.**
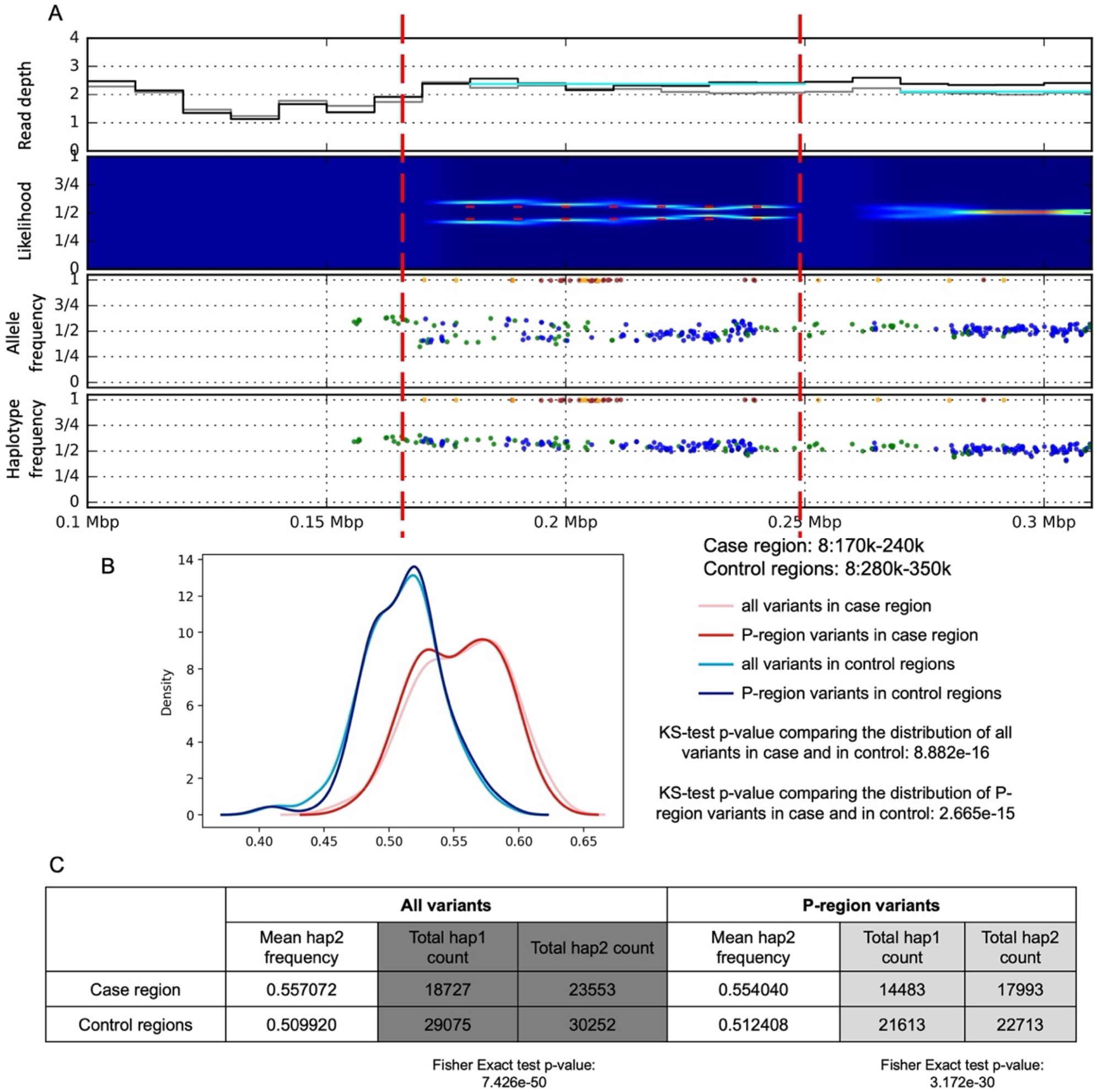
Somatic duplication on chromosome 8 (chr8:170001-240000) identified in. **A)** Read depth, split of likelihood, allele frequency and one-sided haplotype phased allele frequency from bulk data. **B)** Haplotype aware distribution of allele frequencies for SNPs within the predicted somatic duplication region (case, in red) and within upstream and downstream regions (control, in blue). The two distributions are different by KS-test (P-regions are defined by the 1000 Genome Project as accessible to short read sequencing). **C)** Table of read counts for two imputed haplotypes. The imbalance in case compared to control region is significant by Fisher Exact test.

**Figure S17.**
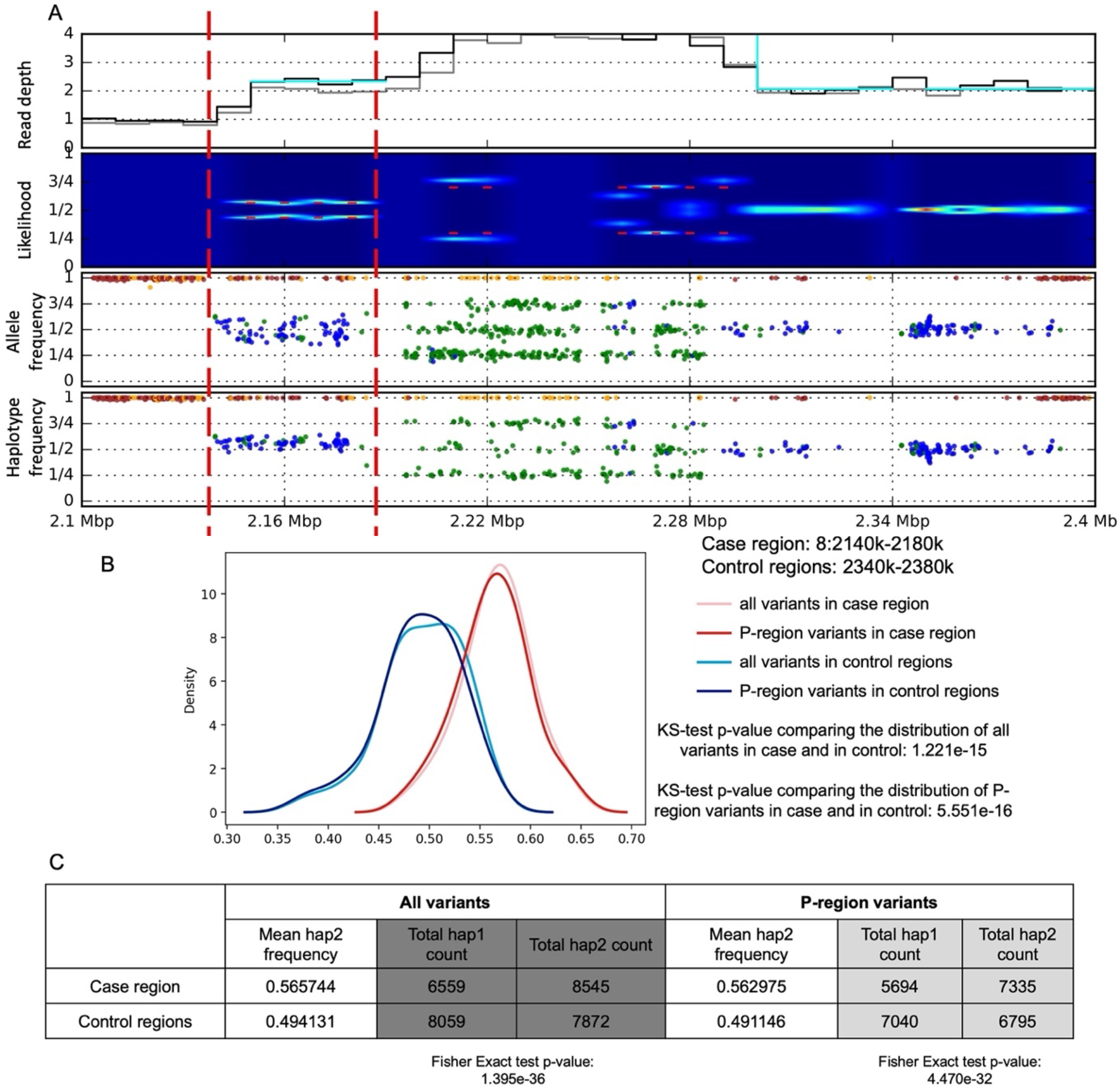
Somatic duplication on chromosome 8 (chr8:2140001-2180000) identified in. **A)** Read depth, split of likelihood, allele frequency and one-sided haplotype phased allele frequency from bulk data. **B)** Haplotype aware distribution of allele frequencies for SNPs within the predicted somatic duplication region (case, in red) and within upstream and downstream regions (control, in blue). The two distributions are different by KS-test (P-regions are defined by the 1000 Genome Project as accessible to short read sequencing). **C)** Table of read counts for two imputed haplotypes. The imbalance in case compared to control region is significant by Fisher Exact test.

**Figure S18.**
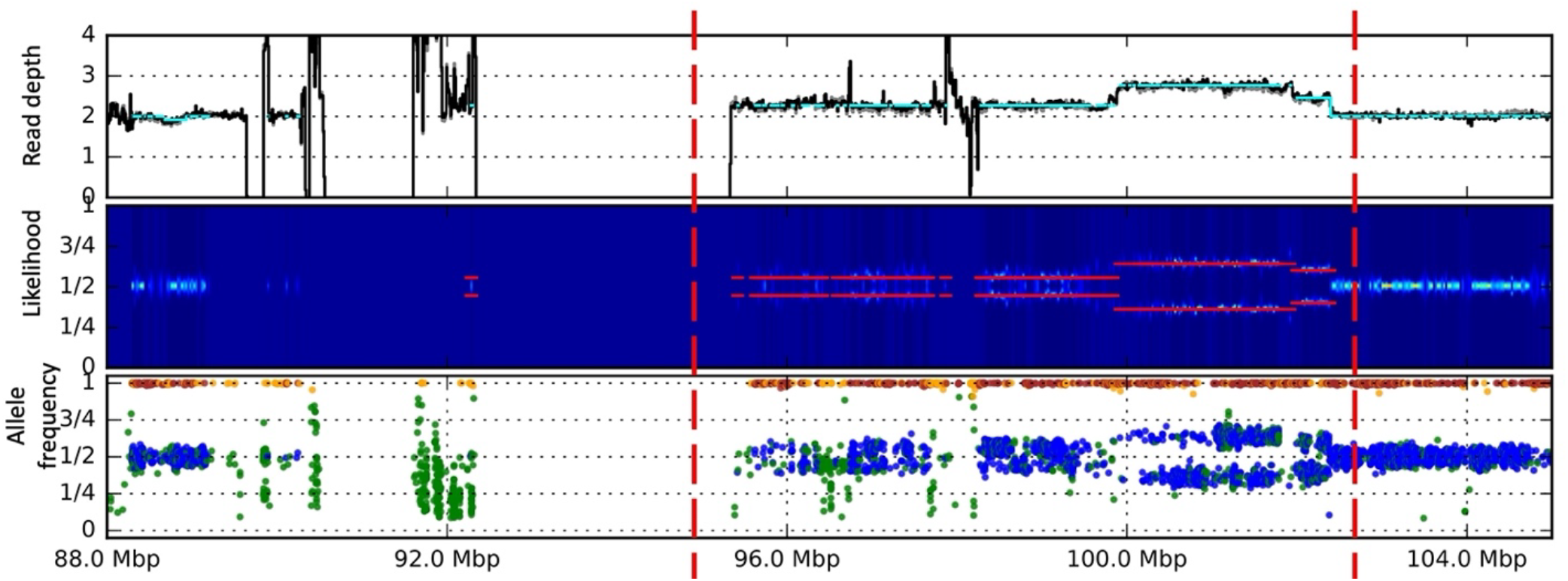
Somatic complex duplication on chromosome 2 (chr2:96200001-102400000) identified from. Increase of read depth, split of likelihood and allele frequency within the complex duplication from cram file of individual.

**Figure S19.**
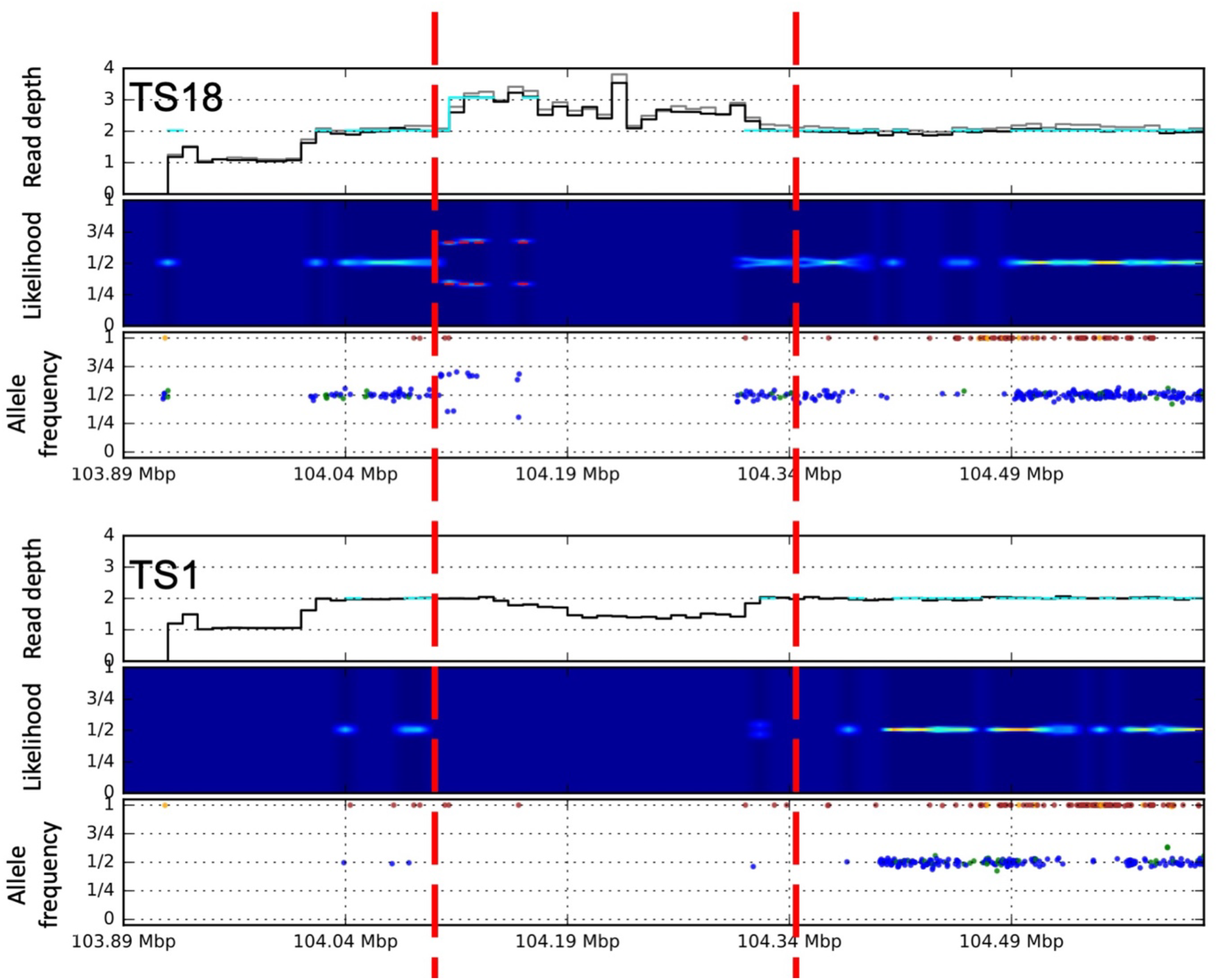
Germline CNV on chromosome 1 (chr1:104100001-104310000) identified in individual within a complex genomic region. at the bottom shows a control without any CNVs within this region.

**Figure S20.**
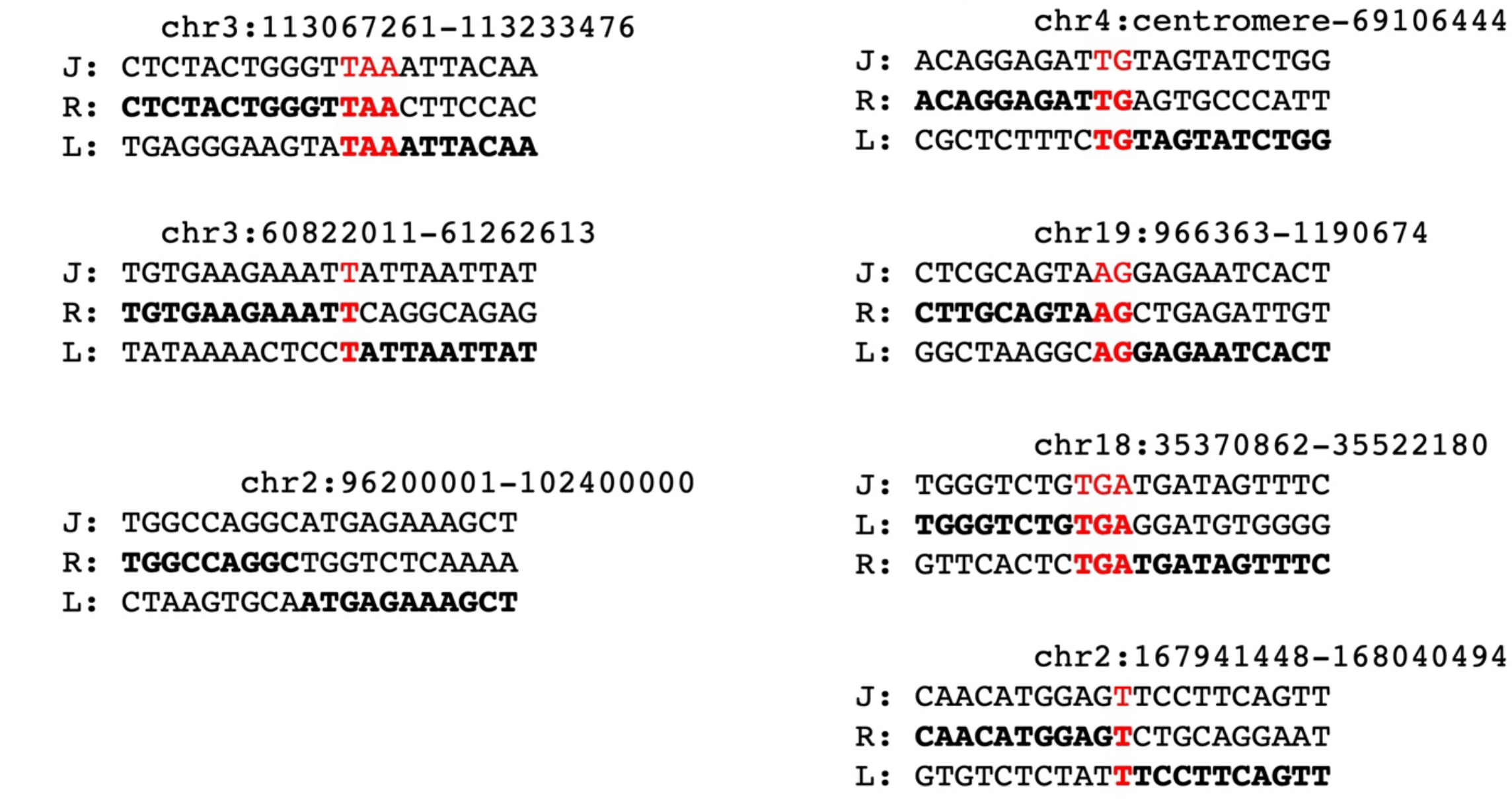
Microhomology for the breakpoints of somatic CNVs identified with specific breakpoints. J stands for the sequence at junction of rearrangements; R stands for the reference sequence at the right breakpoint; L stands for the reference sequence at the left breakpoint.

**Figure S21.**
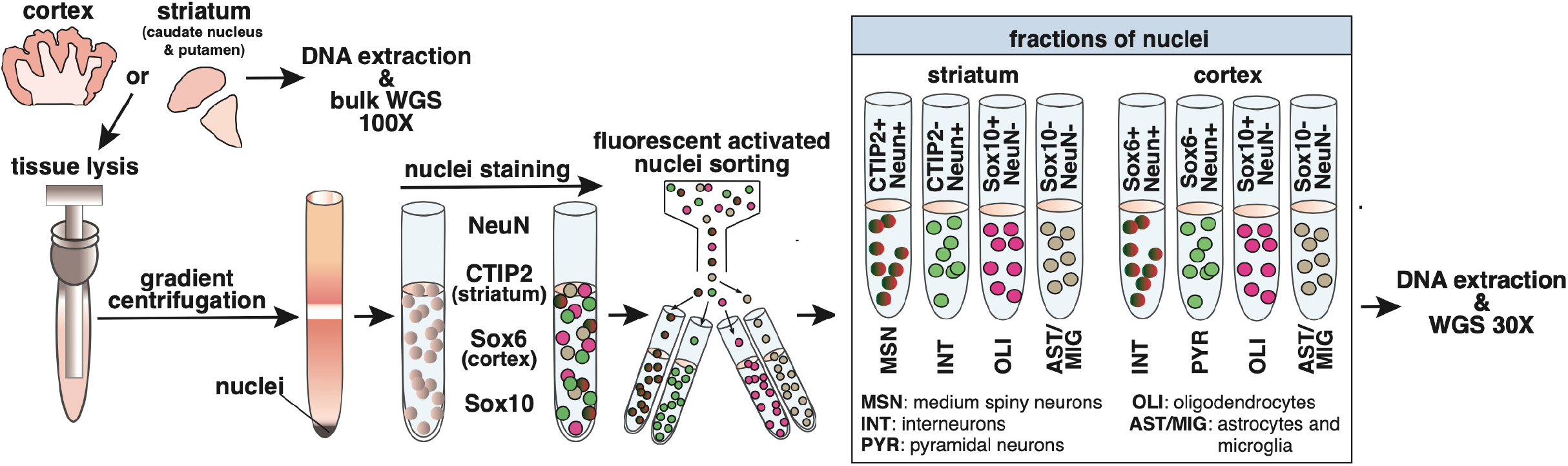
Schematic diagram of fractionation experiments.

**Figure S22.**
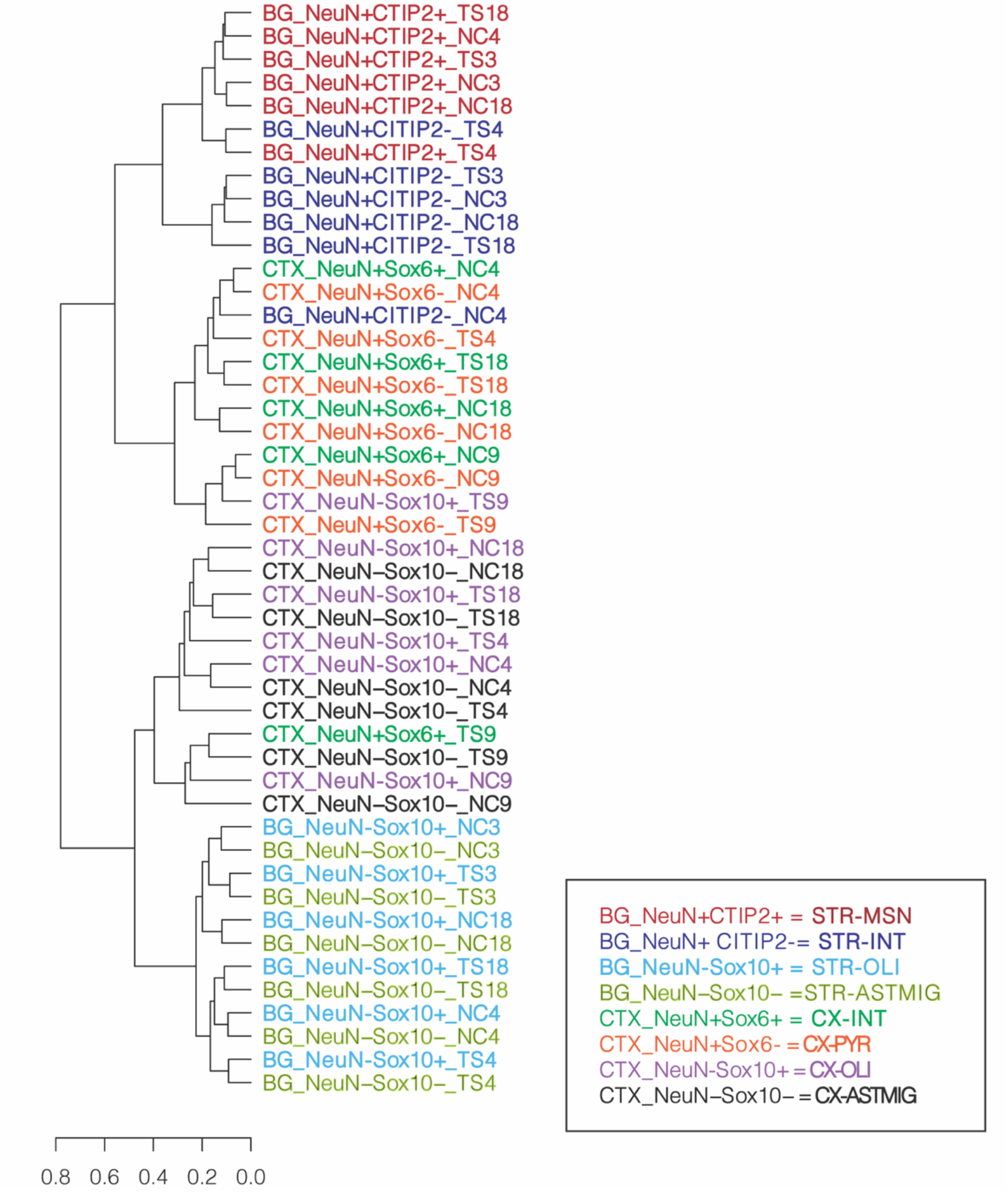
Validation of antibodies used to generate nuclear fractions of TS and matched NC brains by hierarchical clustering based on top 500 variable genes. Hierarchical clustering of transcriptome data shows separation between neurons (NeuN+) and glia (NeuN-) in both cortex (CTX) and striatum (BG), validating the conjugated NeuN-488 antibody. The conjugated CTIP2-647 antibody was validated from clustering of NeuN+/CTIP2+ medium spiny neurons (MSN) and NeuN+/CTIP2-interneurons (INT) in striatum. In contrast the separation of cortical pyramidal neurons (CTX-PYR) (SOX6-NeuN+) and cortical interneurons (CX-INT) (SOX6+NeuN+) via the SOX6 antibody as well as the separation between oligodendrocytes (OLI) versus astrocytes and microglia (ASTMIG) via the SOX10 antibody was not confirmed by the hierarchical clustering.

**Figure S24.**
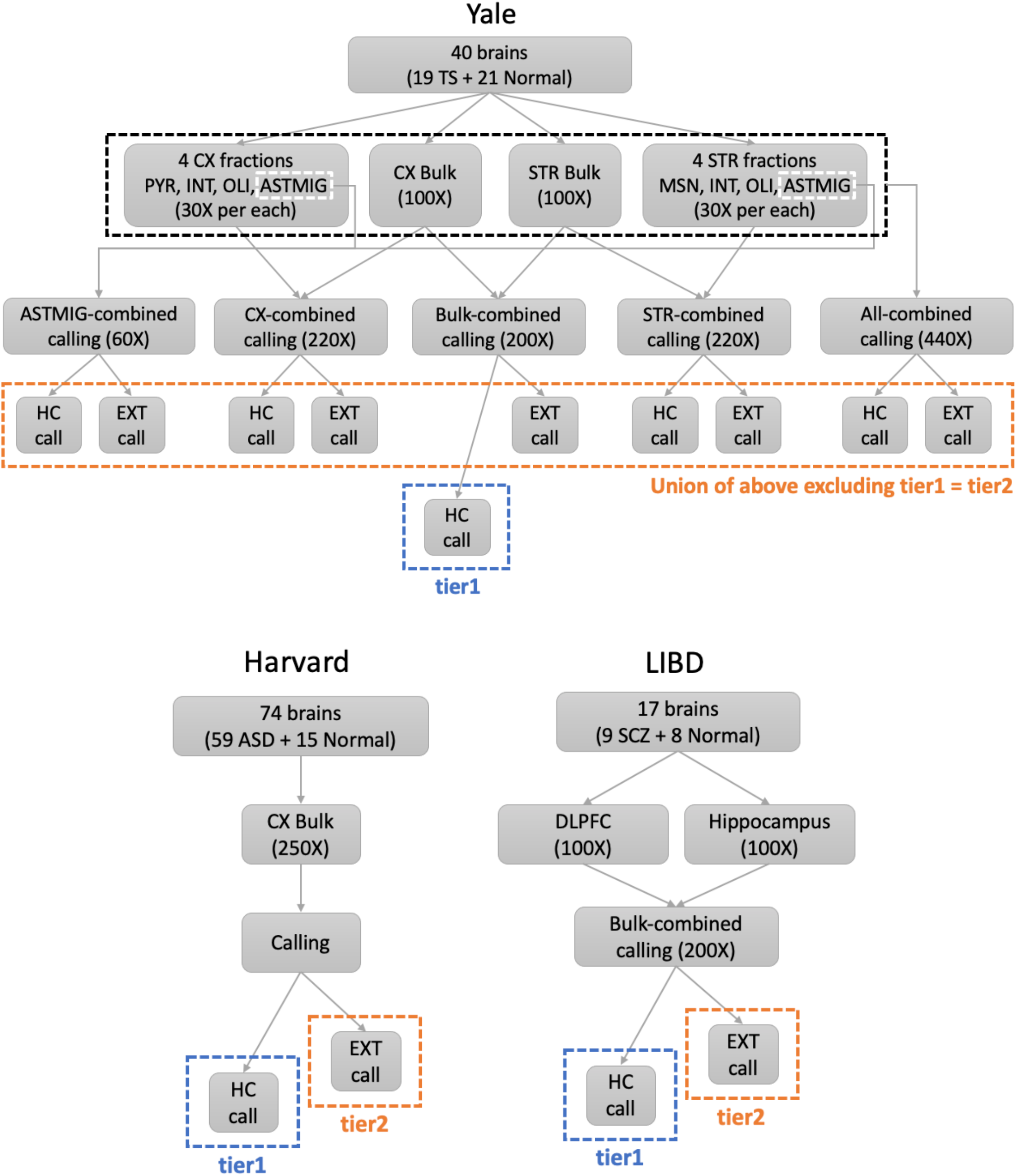
Calling strategy of somatic point mutations per data from each lab (HC: High confidence, EXT: Extended).

**Table S3.** Somatic mutations conferring the gain in TFBS for MEIS1/MEIS2/MEIS3 shown in **Fig. 4C**.

| chromosome | position | reference | alternative | ID |
| --- | --- | --- | --- | --- |
| 5 | 58102459 | A | G | Mut1 |
| 13 | 110724971 | G | A | Mut2 |
| 6 | 43891494 | C | T | Mut3 |
| 17 | 75991384 | C | T | Mut4 |
| 3 | 146341770 | G | A | Mut5 |
| 4 | 1977081 | G | A | Mut6 |
| 14 | 24345503 | G | A | Mut7 |
| 1 | 171052465 | G | A | Mut8 |
| 2 | 179673807 | C | T | Mut9 |
| 22 | 25530565 | G | A | Mut10 |
| 12 | 66341643 | C | T | Mut11 |
| 12 | 92205843 | G | T | Mut12 |
| 12 | 48177099 | G | A | Mut13 |
| 16 | 24642707 | G | A | Mut14 |
| 2 | 129109311 | G | A | Mut15 |
| 14 | 52387789 | G | T | Mut16 |

**Table S4.**
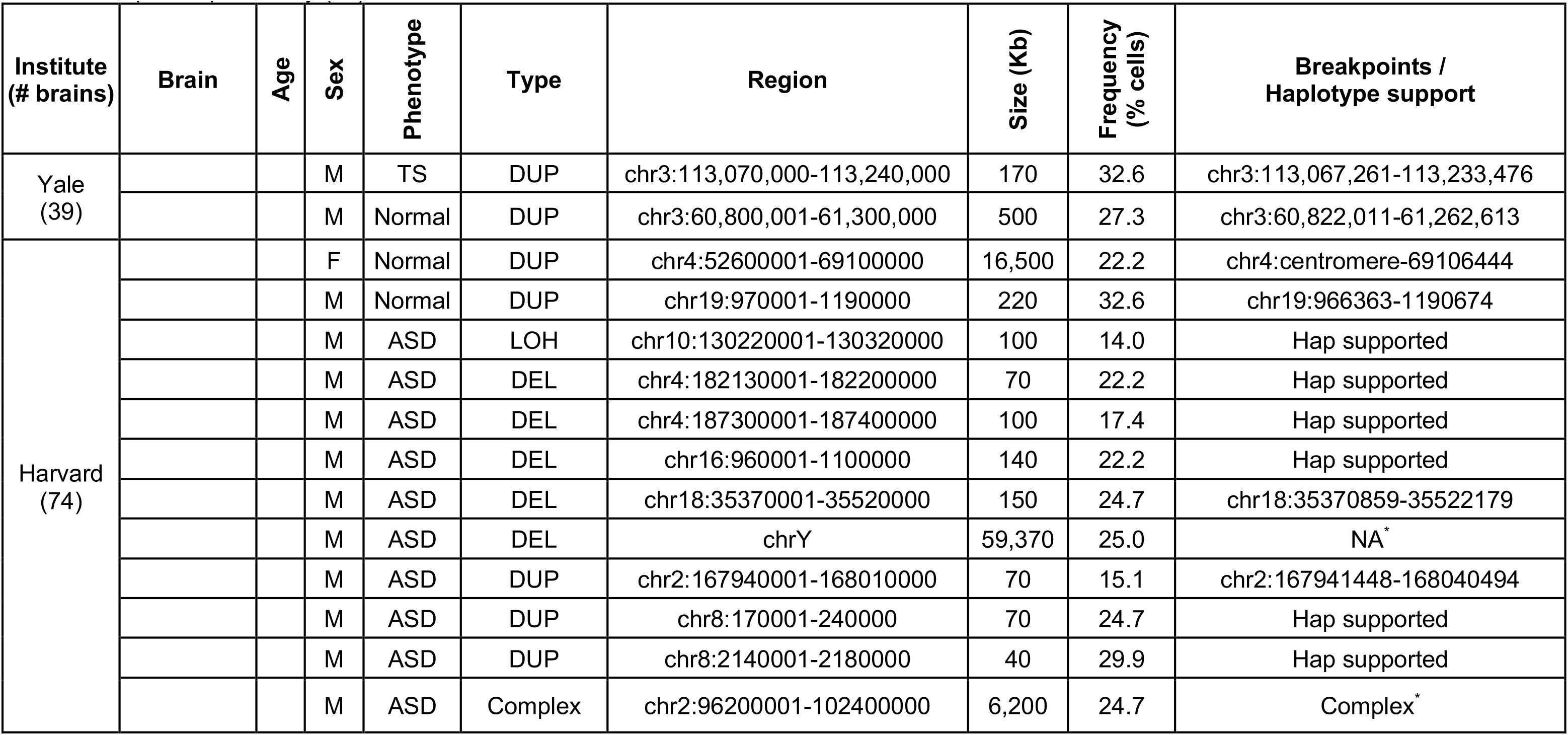
Summary of detected structural mutations. Brains are grouped by the production institute and ordered by phenotype. Regions show boundaries called from application of read depth and BAF analyses. The last column lists, if applicable, precise breakpoints determined with the analysis or paired reads. For events with undetermined breakpoints, evidence for haplotype imbalance was investigated and reported. *structural mutations reported previously (*27*).

**Table S5.**
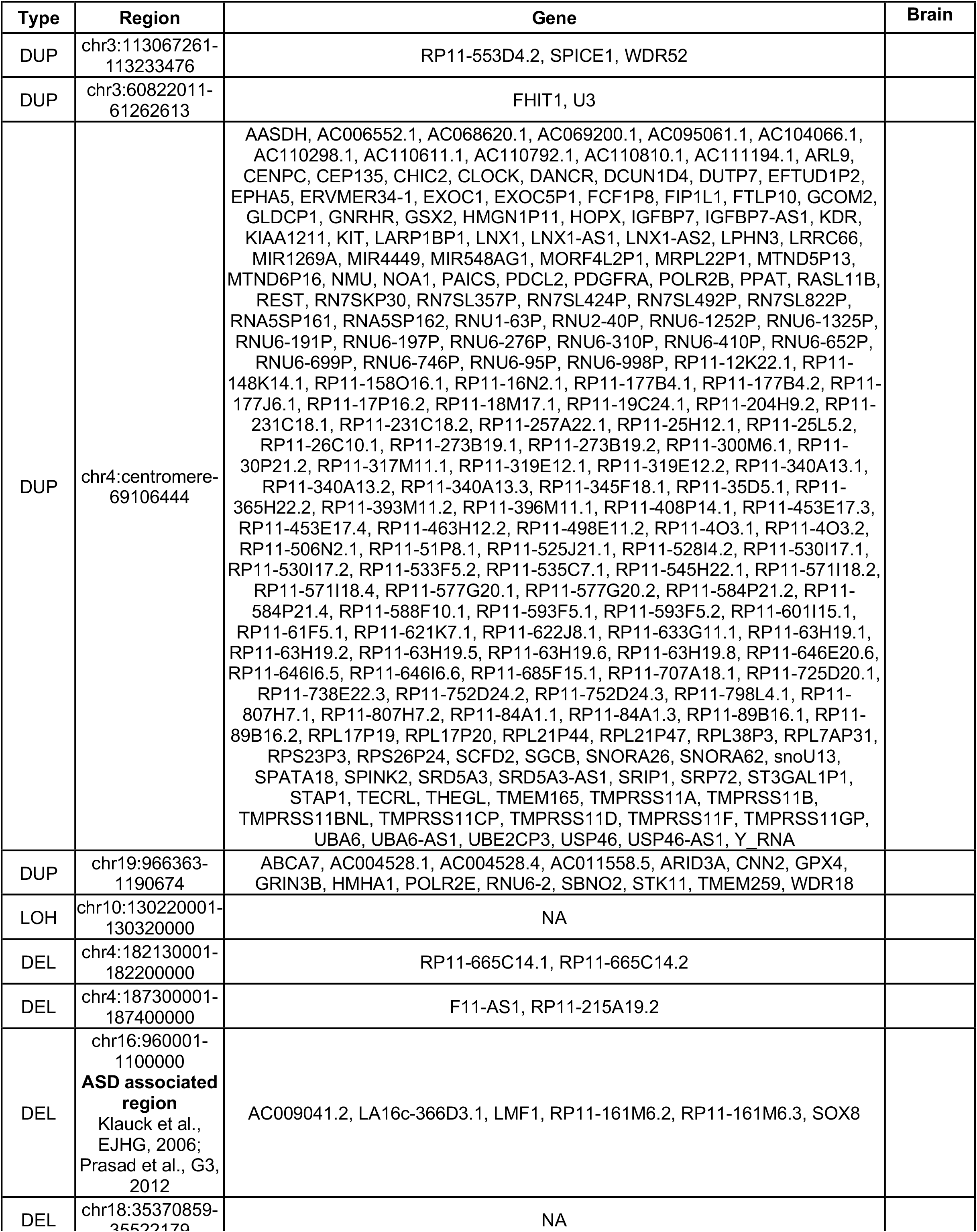
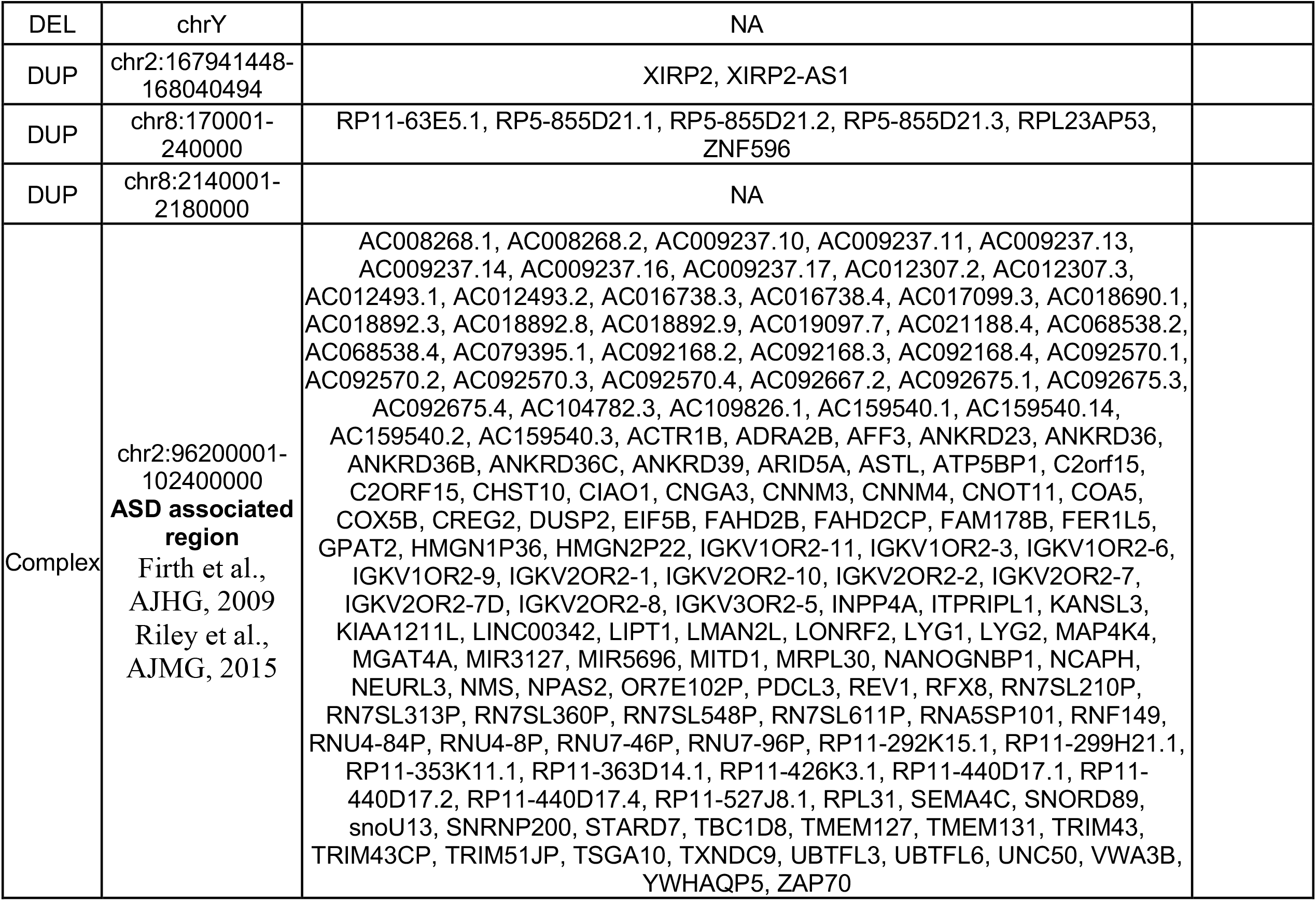
Genes within somatic structural mutations.

## The BSMN consortium

**Executive Committee:** Walsh, Christopher A.; Park, Peter J., Sestan, Nenad; Weinberger, Daniel; Moran, John V.; Gage, Fred H.; Vaccarino, Flora M.; Gleeson, Joseph; Mathern, Gary; Courchesne, Eric; Roy, Subhojit; Chess, Andrew J.; Akbarian, Schahram

**Boston Children’s Hospital:** Sara Bizzotto, Michael Coulter, Caroline Dias, Alissa D’Gama, Javier Ganz, Robert Hill, August Yue Huang, Sattar Khoshkhoo, Sonia Kim, Alice Lee, Michael Lodato, Eduardo A. Maury, Michael Miller, Rebeca Borges-Monroy, Rachel Rodin, Christopher A. Walsh, Zinan Zhou

**Harvard University:** Craig Bohrson, Chong Chu, Isidro Cortes-Ciriano, Yanmei Dou, Alon Galor, Doga Gulhan, Minseok Kwon, Joe Luquette, Peter Park, Maxwell Sherman, Vinay Viswanadham

**Icahn School of Medicine at Mount Sinai:** Schahram Akbarian, Andrew Chess, Attila Jones, Chaggai Rosenbluh

**Kennedy Krieger Institute:** Sean Cho, Ben Langmead, Jeremy Thorpe

**Lieber Institute for Brain Development:** Jennifer Erwin, Andrew Jaffe, Michael McConnell, Rujuta Narurkar, Apua Paquola, Jooheon Shin, Richard Straub, Daniel Weinberger

**Mayo Clinic Rochester:** Alexej Abyzov, Taejeong Bae, Yeongjun Jang, Yifan Wang

**Sage Bionetworks:** Cindy Molitor, Mette Peters

**Salk Institute for Biological Studies:** Fred Gage, Sara Linker, Patrick Reed, Meiyan Wang

**Stanford University:** Alexander Urban, Bo Zhou, Xiaowei Zhu, Reenal Pattni

**Universitat Pompeu Fabra:** Aitor Serres Amero, David Juan, Irene Lobon, Tomas Marques-Bonet, Manuel Solis Moruno, Raquel Garcia Perez, Inna Povolotskaya

**University of Barcelona:** Eduardo Soriano

**University of California, Los Angeles:** Gary Mathern

**University of California, San Diego:** Danny Antaki, Dan Averbuj, Laurel Ball, Martin Breuss, Eric Courchesne, Joseph Gleeson, Xiaoxu Yang, Changuk Chung

**University of Michigan:** Sarah B. Emery, Diane A. Flasch, Jeffrey M. Kidd, Huira C. Kopera, Kenneth Y. Kwan, Ryan E. Mills, John B. Moldovan, John V. Moran, Chen Sun, Xuefang Zhao, Weichen Zhou, Trenton J. Frisbie, Yifan Wang

**Yale University:** Adriana Cherskov, Liana Fasching, Alexandre Jourdon, Sirisha Pochareddy, Soraya Scuderi, Nenad Sestan, Flora M. Vaccarino

